# United States influenza 2022-2023 season characteristics as inferred from wastewater solids, influenza hospitalization and syndromic data

**DOI:** 10.1101/2023.09.12.23295371

**Authors:** Mary E. Schoen, Amanda L. Bidwell, Marlene K. Wolfe, Alexandria B. Boehm

## Abstract

Influenza A virus (IAV) causes significant morbidity and mortality in the United States and has pandemic potential. Identifying IAV epidemic patterns is essential to inform the timing of vaccines and non-pharmaceutical interventions. In a prospective, longitudinal study design, we measured IAV RNA in wastewater settled solids at 163 wastewater treatment plants across 33 states to characterize the 2022-2023 influenza season at the state, health and human services (HHS) regional, and national scales. Influenza season onset, offset, duration, peak, and intensity using IAV RNA in wastewater were compared with those determined using laboratory-confirmed influenza hospitalization rates and outpatient visits for influenza-like illness (ILI). The onset for HHS regions as determined by IAV RNA in wastewater roughly corresponded with those determined using ILI when the annual geometric mean of IAV RNA concentration was used as baseline (i.e., the threshold that triggers onset), although offsets between the two differed. IAV RNA in wastewater provided early warning of onset, compared to the ILI estimate, when the baseline was set at twice the limit of IAV RNA detection in wastewater. Peak when determined by IAV RNA in wastewater generally preceded peak determined by IAV hospitalization rate by two weeks or less. Wastewater settled solids data is an IAV-specific indicator that can be used to augment clinical surveillance for seasonal influenza epidemic timing and intensity.

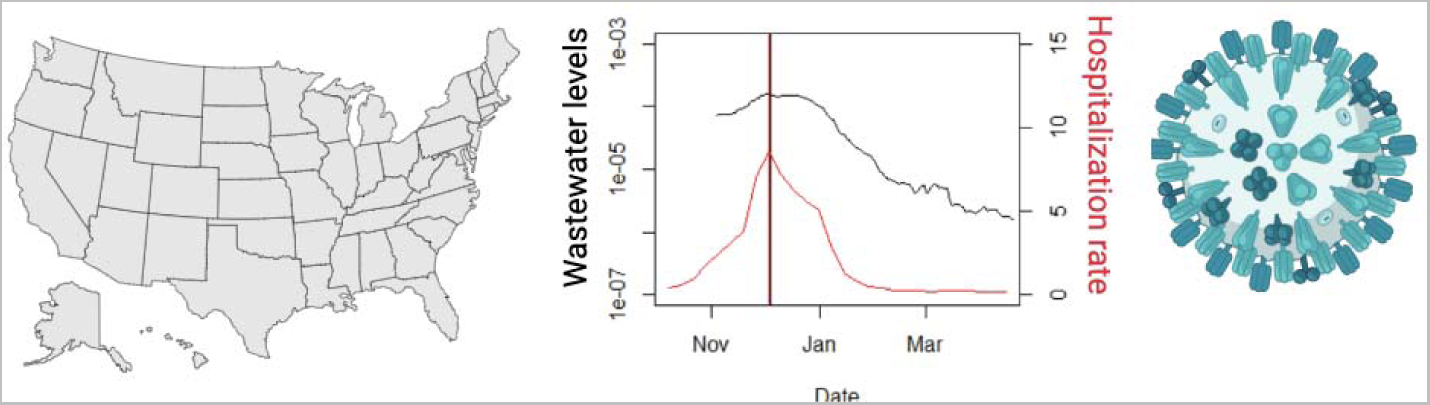

## 1 Introduction

Influenza causes significant morbidity and mortality in the United States (US) causing an estimated 44 million illnesses and upwards of 41,000 deaths annually, and costing $16.3 billion in lost wages and hospitalizations.^1^ Influenza can be caused by influenza A and B viruses; however, influenza B infections have been less common in recent years in the US.^2^ Influenza vaccines are available annually and reduce the risk of severe illness.^1^ However, immune history and antigenic mismatch between the vaccine and circulating strains can modulate vaccine effectiveness.^3^ IAV also has pandemic potential, owing to recombination that occurs frequently among influenza A virus (IAV) subtypes. Several subtypes emerged over the last decade that cause severe illness and evaded seasonal vaccines.^4,5^

Understanding the timing and severity of the annual influenza season is essential in guiding public health recommendations and planning for hospital and clinical resources. The US Center for Disease Control and Prevention (CDC) along with researchers characterize the US influenza season by estimating the onset, offset, duration, peak and intensity (defined in Table 1) using three clinical indicators with historical data: 1) outpatient visits for influenza-like illness (ILI); 2) influenza-related hospitalizations; and 3) influenza- and pneumonia-related deaths.^6–8^ These clinical indicators have been used for decades and allow comparison across influenza seasons, but have known limitations. For example, ILI can include other respiratory illnesses such as COVID-19 and relies on care-seeking behaviors of infected individuals. Hospitalizations and deaths reflect severe illness, and occur well after initial infections occur. Both indicators also have a lag in reporting; are reported at relatively large scales, e.g. aggregated by state or US Health and Human Services (HHS) region; and, in the case of hospitalizations, have limited coverage across the US.^6^ Recently, wastewater concentrations of IAV RNA have been shown to reflect occurrence of influenza A in contributing communities. ^9^ In this study, we investigate how IAV RNA concentrations in wastewater, measured throughout the US in a prospective, longitudinal study, can augment traditional, clinical influenza indicators to characterize the 2022-2023 influenza season.

**Table 1.** Summary of influenza season characteristics, inputs, and approach details. Inputs were limited to those with sufficient available data for wastewater measurements or clinical measures as explained in the methods. NA: not applicable.

| Characteristics | Description | Input | Inclusion Criteria | Approach |
| --- | --- | --- | --- | --- |
| Onset/Offset | Day when measure was above/below the baseline value and remained there for at least two additional weeks | Avg_IAV_Region (R1, R4, R5, R8, and R9) | Regions with collection between 6/1/2022 - 5/31/2023 with coverage of two or more states. | Wastewater baseline values were calculated as described in Section 2.3. Methods 2 and 3 require 11 months of collection. ILI baseline values were obtained from FluView. Baseline values were compared to the input time series to identify onset and offset based on conditions in the description. |
|  |  | Avg_IAV_State (AL, CA, CO, FL, GA, IL, KS, MI, NH, and TX) | States with collection between 6/1/2022 - 5/31/2023 with coverage of two or more WWTPs. |  |
|  |  | ILI Region (R1, R4, R5, R8, and R9) | Regions with available IAV wastewater data. |  |
| Duration | The number of days between onset and offset | Same as Onset/Offset. |  |  |
| Peak/Intensity | The date/magnitude of the maximum input value during an influenza season | Avg_IAV_State (CA, CO, GA, MN, MI, and UT) | States with hospitalization data and coverage of two or more WWTPs between 6/1/2022 and 5/30/2023. | The maximum of the input time series was identified to characterize peak and intensity. |
|  |  | State hospitalization rate (CA, CO, GA, MN, MI, and UT) | States with available IAV wastewater data. |  |
|  |  | Avg_IAV_US | NA |  |
|  |  | Overall hospitalization rate | NA |  |

To our knowledge, several previous studies have compared IAV RNA in wastewater to influenza occurrence in communities contributing to wastewater. Wolfe et al.^9^ showed IAV RNA was enriched in wastewater solids compared to liquid wastewater and that concentrations correlated well with cases identified during active influenza surveillance efforts on two university campuses. Mercier et al.^10^ used IAV concentrations in wastewater and sludge to forecast a municipal IAV outbreak and provide real-time information on the outbreak subtype. Stadler et al.^11^ showed the presence of IAV RNA in liquid wastewater correlated to the presence of IAV at K-12 schools. Boehm et al.^12^ showed IAV RNA in wastewaters solids at a large plant serving 1.5 million people correlated with state-aggregated IAV test positivity rates. Lastly, Boehm et al.^13^ identified a “tripledemic” when concentrations of IAV, respiratory syncytial virus (RSV), and SARS-CoV-2 RNA in wastewater solids peaked at approximately the same time in the Greater San Francisco Bay Area of California; the wastewater data was used to identify localized onset and offset of wastewater events. These previous studies have focused on small geographical areas. This study aims to expand on these to examine IAV RNA concentrations in wastewater across the entire US.

This work compares onset, offset, duration, peak, and intensity of the 2022-2023 influenza season using clinical data and IAV RNA concentrations in wastewater settled solids aggregated to match the spatial scales over which clinical data are typically aggregated. Influenza season characteristics are then estimated at a smaller scale using IAV concentrations to illustrate the progression of onset.

## 2.0 Methods

### 2.1 Input data

#### 2.1.1 Clinical data

Influenza hospitalization rates (hereafter hospitalization rates) are calculated as the number of residents of a defined area who are hospitalized with a positive influenza laboratory test divided by the total population within the area. State hospitalization rates are calculated and reported by the Influenza Hospitalization Surveillance Network (FluSurv-NET), a collaboration between CDC, the Emerging Infections Program Network, and state and local health departments in 13 geographically distributed areas in the US that conduct population-based surveillance.^14^ FluSurv-NET hospitalization rates are reported on a weekly basis, anchored to the Saturday marking the end of each week of the year, and organized by influenza season. Hospitalization rates in units of hospitalizations per 100,000 population were downloaded from FluSurv-NET for all available states for the 2022-2023 influenza season including CA, CO, CT, GA, MD, MI, NM, NY, OH, OR, TN, and UT along with an overall network rate (herein referred to as the overall hospitalization rate).

The US Outpatient Influenza-like Illness Surveillance Network (ILINet) collects information on outpatient visits from 2000 out-patient health-care providers in all 50 states for respiratory illness.^15^ ILINet captures visits due to any respiratory pathogen that presents with the symptoms of fever plus cough or sore throat. The percentage of patient visits to healthcare providers for ILI for the US, HHS regions, and states is reported by influenza season on a weekly basis, anchored to the Saturday marking the end of each week of the year, through the online FluView Interactive platform.^2^

To identify onset and offset, CDC calculates a ILI “baseline level” during non-flu weeks for HHS regions 1-10 (Figure S1) and the US. The baselines are developed by calculating the mean ILI during non-influenza weeks for the most recent three seasons excluding the COVID-19 pandemic and adding two standard deviations.^15^ We viewed the baseline levels along with the timeseries of ILI for HHS regions in FluView Interactive.

#### 2.1.2 IAV RNA in wastewater solids

Samples were collected typically three times per week at up to 163 wastewater treatment plants (WWTPs) between 5 January 2022 and 31 May 2023 (Figure S2 and Table S1). Maximum sampling frequency was daily at 8 CA WWTPs. The date on which WWTPs started participating in the prospective monitoring effort varied between 5 January 2022 and 24 May 2023 (Table S1).

Samples of settled solids were collected from the primary clarifier, or solids were obtained from raw influent by either using an Imhoff cone^16^, or allowing the influent to settle for 10-15 mins, and using a serological pipette to aspirate the settled solids into a falcon tube (Table S1).

Samples were collected by WWTP staff and sent at 4°C to the laboratory where they were processed immediately. The time between sample collection and receipt at the lab was typically between 1-3 days; during this time limited degradation of the RNA targets is expected.^17,18^ Table S1 provides additional information on the WWTPs including the type of sample collected from the WWTP, the populations served and number of samples collected. In total, these WWTPs serve 11.6% of the US population. A total of 18,590 samples were collected and analyzed.

Wastewater solids were subjected to pre-analytical processing to obtain nucleic-acid extracts as reported in detail in a number of different publications ^12,19,20^ and on protocols.io and outlined briefly in the supporting information (SI). Bovine coronavirus (BCoV) vaccine was spiked into each sample to gain insight into nucleic-acid recovery. The nucleic-acids were then used as templates in droplet digital RT-PCR reactions to measure concentrations of the IAV M1 gene ^21^, pepper mild mottle virus (PMMoV) RNA^21^, and BCoV RNA^21^. The analytical approaches including the thresholding of the fluorescent values from the instrument are described in detail in the data descriptor by Boehm et al.^21^ and are not repeated here. Concentrations are reported in units of copies per gram dry weight (cp/g). Positive and negative extraction and RT-PCR controls were run alongside all samples, as described elsewhere^21^ and in the SI. These data have not been previously published aside from samples collected between 7/1/22 and 5/7/23 at eight of the 163 WWTPs in this study, all eight are located in the greater San Francisco Bay Area of California.^13^

### 2.2 Data analysis

#### 2.2.1 Aggregated IAV/PMMoV

We aggregated the wastewater data across WWTPs to calculate daily average concentrations of IAV RNA normalized by PMMoV RNA for states and HHS regions. PMMoV normalization was done to account for potential differential recoveries of viral nucleic acids during the pre-analytical steps,^22^ and because a mass balance models suggest that IAV/PMMoV should scale as disease incidence rate.^23^ The Avg_IAV_State_d_ or Avg_IAV_Region_d_ is the population weighted average of IAV/PMMoV_n,d_ on day *d* from *N* WWTPs contributing to the state or region calculated as:

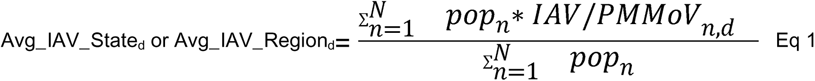

where pop_n_ is the population served by WWTP *n*. IAV/PMMoV_n,d_ is the 5-sample trimmed average of IAV/PMMoV from plant *n* on day *d*. Prior to calculating the aggregated line, non-detect values are set to the limit of detection of 500 cp/g normalized by the average PMMoV value for the WWTP. For days when a WWTP did not have a sample, IAV/PMMoV_n,d_ was estimated using linear interpolation between adjacent values. Weekly median Avg_IAV_State or Avg_IAV_Region was calculated (Sunday through Saturday) for comparison with FluSurv-NET hospitalization rate data.

We aggregated the average concentrations across states to calculate daily average concentration of IAV/PMMoV for the US. The Avg_IAV_US_d_ is the population weighted average of Avg_IAV_State_s,d_ on day *d* from *S* states calculated as:

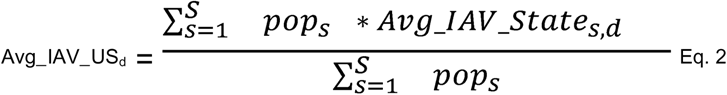

where pop_s_ is the population of state *s* and Avg_IAV_State_s,d_ is calculated for state *s* using equation 1. We aggregated over state, as opposed to WWTPs, to avoid oversampling of states with many participating WWTPs. Only states with coverage of two or more WWTPs at time of analysis were included in Eq. 2 (i.e., AL, CA, CO, FL, GA, ID, IL, IN, IA, KS, ME, MD, MI, MN, NH, NJ, NC, OH, PA, TX, UT, VT, and VA; abbreviation in Table S1). Weekly median Avg_IAV_US_d_ was calculated (Sunday through Saturday) for comparison with overall hospitalization rate data.

#### 2.2.2 Influenza Season Characteristics

Table 1 summarizes the approaches used to estimate the influenza season characteristics of onset, offset, duration, peak, and intensity for each data input. Influenza surveillance by CDC begins 10/2/2022, and ends 9/30/2023 for the 2022-2023 season; whereas, the period of analysis for this paper was 6/1/2022 - 5/31/2023 to capture the onset in states and regions that occurred earlier than the CDC starting date of 10/2/2022.

For analysis of peak and intensity, we compared characteristics using wastewater to those using hospitalization rates, given that the hospitalization rates are specific to influenza A infections. The comparison was limited to the FluSurv-NET reporting spatial scales of state and the overall network estimate (listed in Table 1). For analysis of onset, offset, and duration, we compared characteristics using wastewater to those using ILI, given that ILI is used by CDC to inform these characteristics.^15^ The comparison was limited to the ILI reporting scale of HHS region (listed in Table 1). Onset, offset, and duration were also estimated for states using wastewater data.

#### 2.2.3 Calculation of baseline using wastewater IAV/PMMoV

In an effort to mimic the approaches used by CDC and other researchers in defining influenza season onset using an off-season baseline value,^24^ we calculated wastewater off-season baseline (hereafter “baseline”) values using four different approaches: (1) recording the Avg_IAV_Region values that correspond with the ILI onset and offset dates, (2) calculating the annual Avg_IAV_Region mean over of the period of analysis, (3) calculating the annual Avg_IAV_Region geometric mean over the period of analysis, and (4) calculating twice the minimum Avg_IAV_Region from start of collection (which varied) to the end of the period of analysis (representing roughly twice the lowest detectable concentration). Method 1 is dependent on ILI and interprets the ILI baseline in terms of Avg_IAV_Region. State baseline values were estimated using Avg_IAV_State data and methods 2 - 4; ILI baseline values are not available at the state level. Methods 2 and 3 are based on WHO guidance for the aggregate average method ^25^, but using only one year of data rather than multiple years (as multiple years were not available); while, method 4 does not require a full year of observation. The motivation to use twice the minimum was that during periods devoid of influenza activity, concentrations of IAV RNA in wastewater tend to be non-detect (see results).

#### 2.2.4 Additional analysis

Cumulative empirical curves of the proportion of states with onset during the 2022 - 2023 influenza season were generated using wastewater IAV baselines calculated by methods 2, 3, and 4. Linear relationships were evaluated between log_10_ transformed a) weekly median Avg_IAV_State and hospitalization rate at the state level and b) weekly median Avg_IAV_US and overall FluNet hospitalization rate (details provided in SI).

## 3 Results

### 3.1 QA/QC

The positive and negative controls were positive and negative, respectfully, indicating no contamination. The median BCoV recovery across all the samples was 1.21 (interquartile range = 0.84 to 1.42) suggesting reasonable recovery; recovery values greater than 1 are likely a result of uncertainties associated with quantifying the quantity spiked into the samples. The reporting table from the Environmental Microbiology Minimal Information (EMMI) guidelines has been provided previously in Boehm et al.^21^. Previous work indicated minimal inhibition of the IAV assay using the applied pre-analytical and analytical approaches.^21^

### 3.2 IAV Wastewater Data

Wastewater IAV concentrations collected from 163 WWTPs covering 33 states ranged between below the limit of detection and 15,438,760 cp/g over the period of analysis of 6/1/2022 - 5/31/2023 (Figure S3 and Table S1). The IAV concentrations were normalized by PMMoV (Figure S4) and aggregated to calculate state and HHS regional average concentrations (i.e., Avg_IAV_State and Avg_IAV_Region). The WWTP, state, and HHS regional wastewater concentrations followed seasonal patterns over the period of analysis with elevated concentrations in the winter season compared to summer, but also indicated sporadic increases in concentrations throughout the summer of the 2021-2022 season (e.g., Figures S3 and S4, Figures 1 and 2).

**Figure 1.**
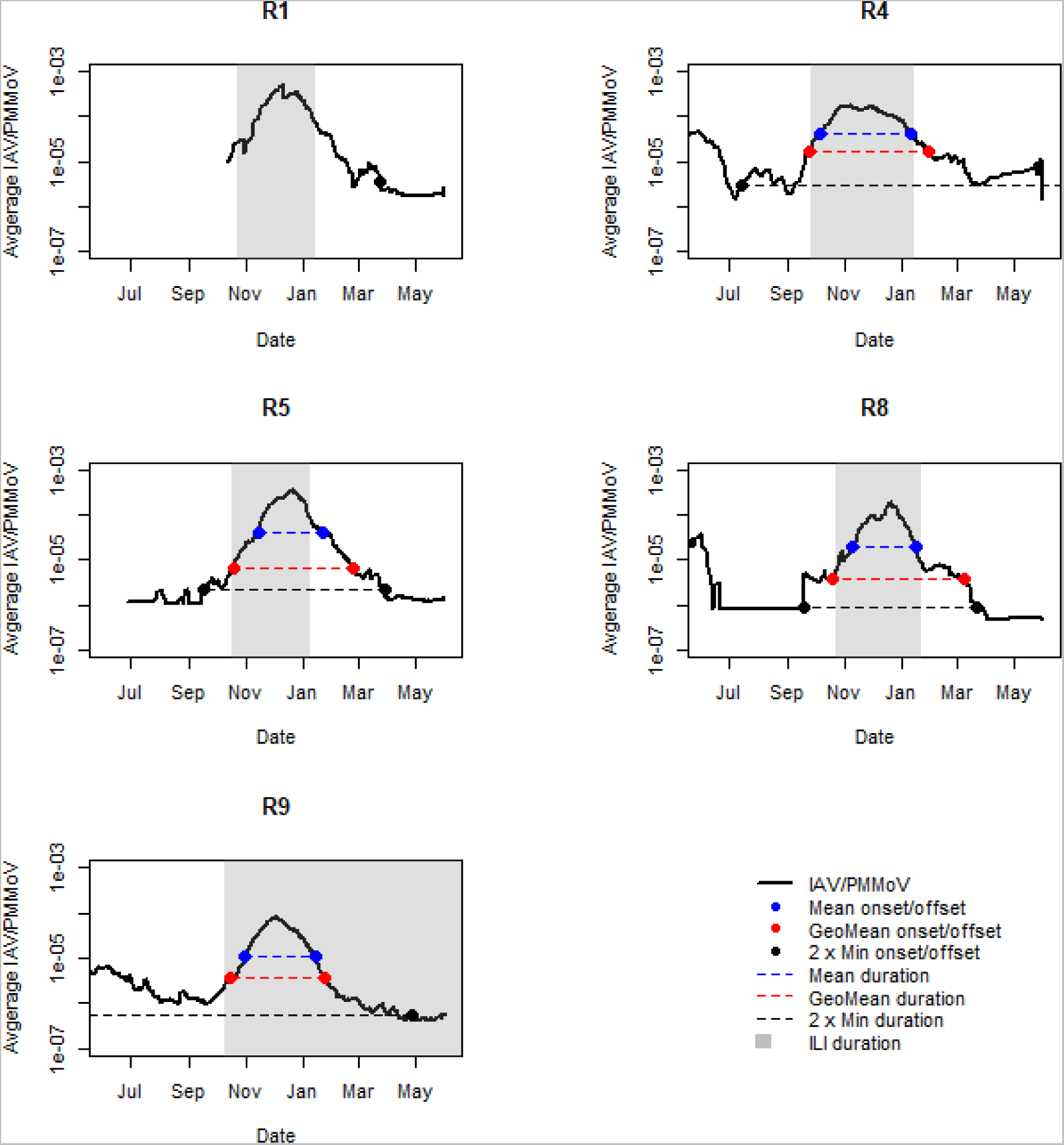
Comparison of alternative baseline Avg_IAV_Region values (y-axis) and corresponding influenza season onset, offset, and duration for HHS regions (abbreviated R) for 2022-2023 influenza season (x-axis). Baseline values were calculated using Avg_IAV_Region annual mean (Mean), annual geometric mean (GeoMean), and twice the minimum observation (2 × Min). Shading indicates ILI onset, offset, and duration. Lines or shading that extend beyond the plotted time period indicate that onset or offset did not occur during the period of analysis using the selected baseline. Missing duration lines and onset/offset points indicate that data was not available for calculation. A map of HHS regions is available Fig S1.

**Figure 2.**
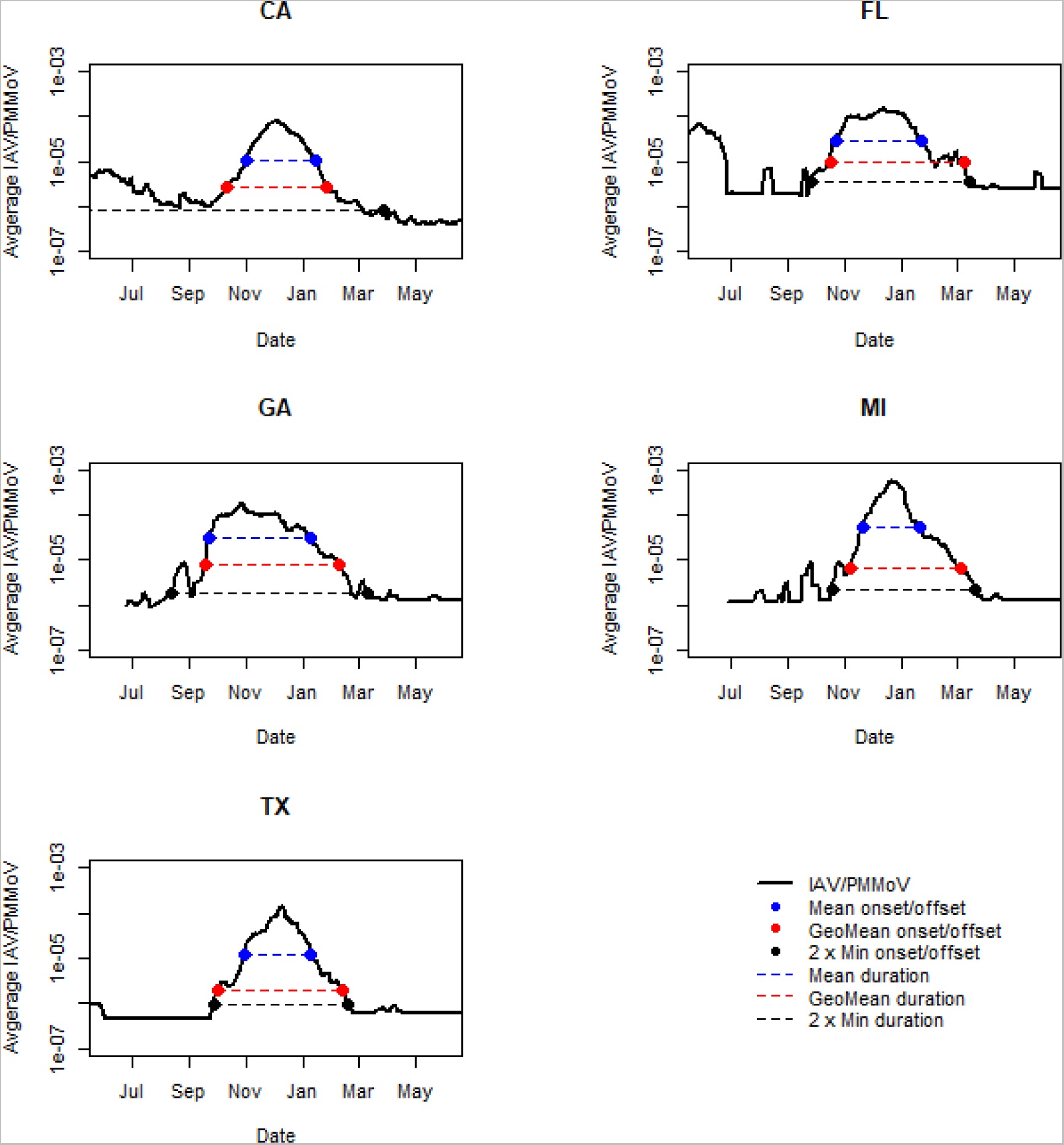
Comparison of alternative baseline Avg_IAV_State values (y-axis) and corresponding influenza season onset, offset, and duration for states with at least 11 months of data for 2022-2023 influenza season (x-axis). Baseline values were calculated using Avg_IAV_State annual mean (Mean), annual geometric mean (GeoMean), and twice the minimum observation (2 × Min). Lines that extend beyond the plotted time period indicate that onset or offset did not occur during the period of analysis using the selected baseline.

### 3.3 IAV baseline values

To identify the onset and offset of influenza season using wastewater, we identified the dates on which the Avg_IAV_State or Avg_IAV_Region exceeded a wastewater baseline value defined in one of four ways described in methods: concentrations corresponding to ILI onset and offset (method 1), the annual mean (method 2), the annual geometric mean (method 3), and twice the minimum (method 4) (Figure 1 and Table S1 for HHS regions, Figure 2 Table S3 for states).

Across states and HHS regions, the lowest wastewater baselines were those defined as twice the minimum (range of 5.70 × 10^−7^ − 3.46 × 10^−6^ across HHS regions and 8.10 × 10^−7^ − 3.54 × 10^−6^ across states) while the highest baselines were those defined as the annual mean (range of 1.06 × 10^−5^ − 4.14 × 10^−5^ across HHS regions and 1.02 × 10^−5^ − 5.12 × 10^−5^ across states), with the exception of HHS region 5 (R5) for which the baseline corresponding to the ILI offset was higher than the annual mean baseline. Baselines defined as the annual geometric mean and corresponding to ILI onset and offset (only applicable to HHS regions) were generally between those described above.

### 3.4 Onset, offset, and duration

#### 3.4.1 HHS Region comparison

The onset, offset and durations of the 2022-2023 influenza season determined using the ILI baseline and wastewater baseline values using methods 2-4 are compared in Figure 1 and Table S4 for HHS regions. Onset was earliest in R4 (Southeast) followed by R9 (West), R5 (Midwest) or R8 (Central), and R1 (Northeast) for all baseline approaches. Note that we did not have sufficient data to examine onsets in the other HHS regions.

Comparing onset determined using ILI and wastewater, onset using the Avg_IAV_Region baseline corresponding to twice the minimum was up to 85 days earlier than onset calculated by the other approaches (range of 30 to 85 days); whereas, the ILI, Avg_IAV_Region annual mean, and Avg_IAV_Region annual geometric mean onset values differed less than 30 days across the HHS regions. Onset determined by the Avg_IAV_Region annual mean was the latest across approaches. Onset for R9 could not be determined using the baseline corresponding to twice the minimum because observations remained elevated throughout all of 2022.

The offset based on the Avg_IAV_Region annual mean baseline fell within a 14 day range (range of 1/8/2023 - 1/22/2023) for HHS regions with available wastewater data. The ILI offset occurred during the same 14 day range, with the exception of R9 which did not reach offset by the end of the analysis (5/31/2023) using ILI baseline. The offset using baselines based on Avg_IAV_Region geometric mean and twice the minimum generally occurred within a wider range (between 1/24/2023 and 4/28/2023). As a result, the influenza season durations were greater using these two approaches rather than ILI baseline or Avg_IAV_Region annual mean baseline (with the exception of R9).

#### 3.4.2 State estimates

State level onset, offset and duration were calculated using Avg_IAV_State annual mean, annual geometric mean, and twice the minimum (Figures 2 and Table S5). The order of onset and offset by method using state data mirrored the HHS region analysis as did the comparison of the duration. Similar to the regional analysis, onset for CA (in R9) could not be determined using the baseline corresponding to twice the minimum because observations remained elevated throughout the period of analysis.

Cumulative curves of the proportion of states achieving onset over time during the 2022-2023 influenza season are compared using baselines of Avg_IAV_Region annual mean (5 states with annual data), annual geometric mean (5 states with annual data) or twice the minimum (9 states with data) (Figure 3). The curves using the baselines of annual geometric mean and annual mean were shifted to the right compared to twice the minimum consistent with their tendency to indicate later onset (also illustrated in Figure 2).

**Figure 3.**
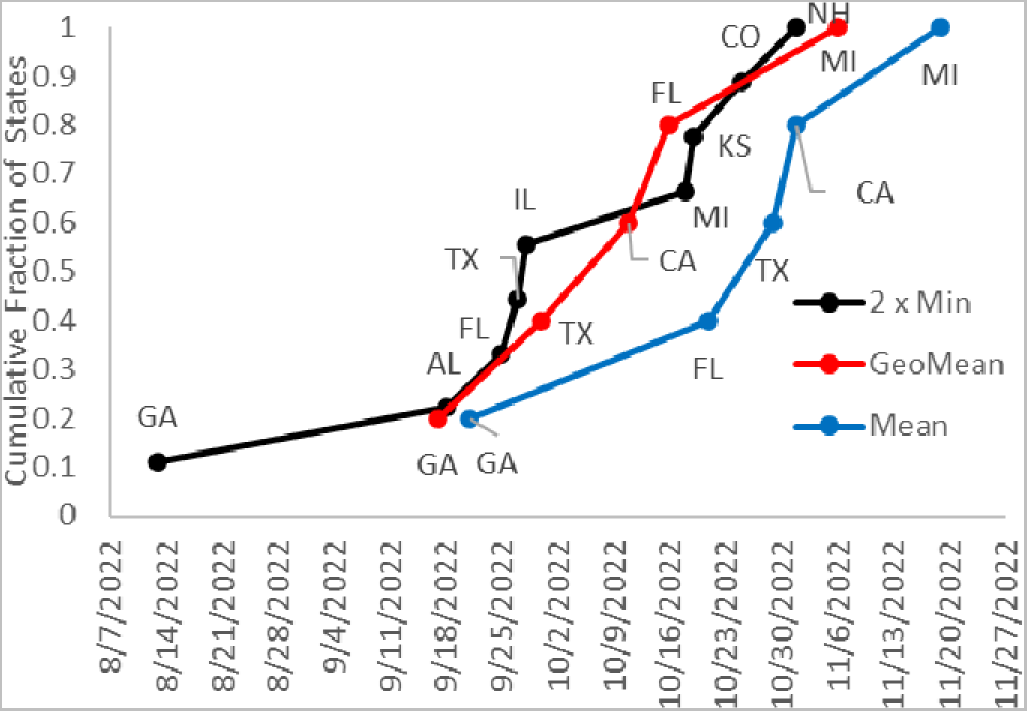
Cumulative curves of the proportion of states with onset (y-axis) over time (x-axis) during 2022-2023 influenza season using Avg_IAV_State baselines of the annual mean (N=5), geometric mean (N = 5) or twice the minimum observation (2 × Min) (N = 9).

The curve using the Avg_IAV_State baseline of twice the minimum was roughly sigmoidal, indicating that onset happened over a narrow time interval for the majority of states despite the earliest onset in GA. GA preceded other states by 36, 30, and 13 days using baselines of twice the minimum, annual geometric mean, and annual mean, respectively. The limited number of states with sufficient data for the mean and geomean baselines made it difficult to assess curve shape. The trajectory of onset moved outward from the origin in the Southeast using all selected baselines. However, the order of onset varied by baseline method; for example, onset in FL occurred before TX using the annual mean and twice the minimum baselines but after using the annual geometric mean baseline.

### 3.5 Peak and Intensity

#### 3.5.1 State comparison

The 2022-2023 influenza season state peaks (i.e., date of highest measure) and intensities (i.e., value at time of peak) are compared using wastewater (Avg_IAV_State) and laboratory-confirmed influenza hospitalizations collected by CDC (hospitalization rate) in Figure 4 and Table S6. The Avg_IAV_State peaks generally preceded the hospitalization rate peaks (range of 2 - 12 days), with the exception of MN. When the intensity as measured by hospitalization rate increased across states, the intensity measured by Avg_IAV_State concentration also increased across states (Table S6). The Avg_IAV_State peak values were roughly two orders of magnitude greater than the Avg_IAV_State baseline values calculated using twice the minimum observation and between roughly 1 and 2 orders of magnitudes greater than the Avg_IAV_State annual mean or annual geometric mean baselines.

**Figure 4.**
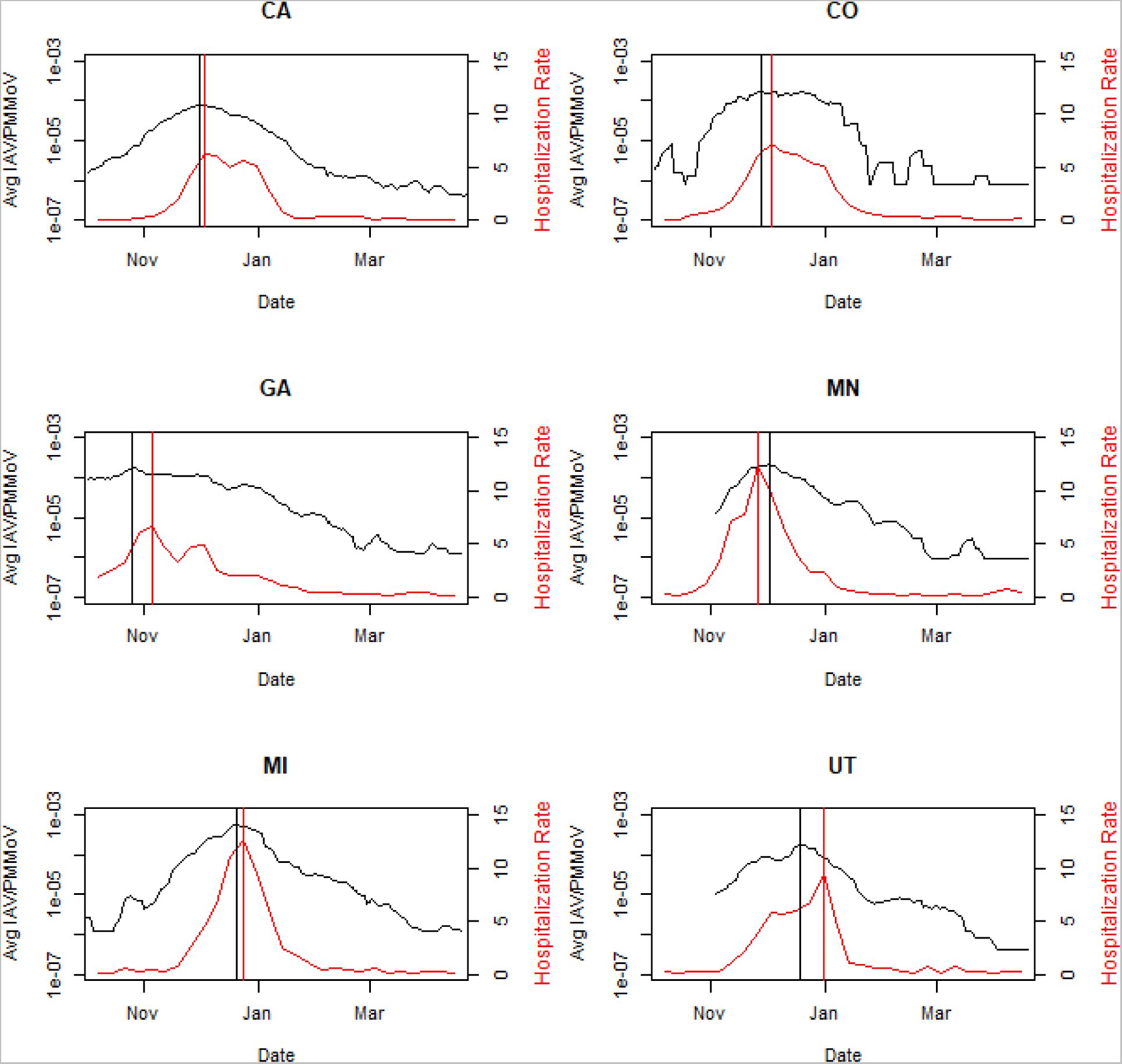
2022-2023 influenza season peak (vertical line) and intensity (y-axis) by state using Avg_IAV_State or FluSurv-NET hospitalization rate (hospitalizations per 100,000 population) for states with both sets of data

#### 3.5.2 US comparison

When Avg_IAV_State were aggregated to create a population weighted US value, the peaks estimated using the Avg_IAV_US and overall hospitalization rate fall one day apart (Figure 5b, 12/03/2022 and 12/04/2022) despite having different representations of states (Figures S5 and S6). The wastewater dataset did not include state data for states with the highest intensities as measured by FluServe-Net (e.g., CT, NM, NY, OR, and TN). FluServe-Net did not include 17 states with wastewater data, particularly those with early season onset (e.g., FL and TX).

**Figure 5.**
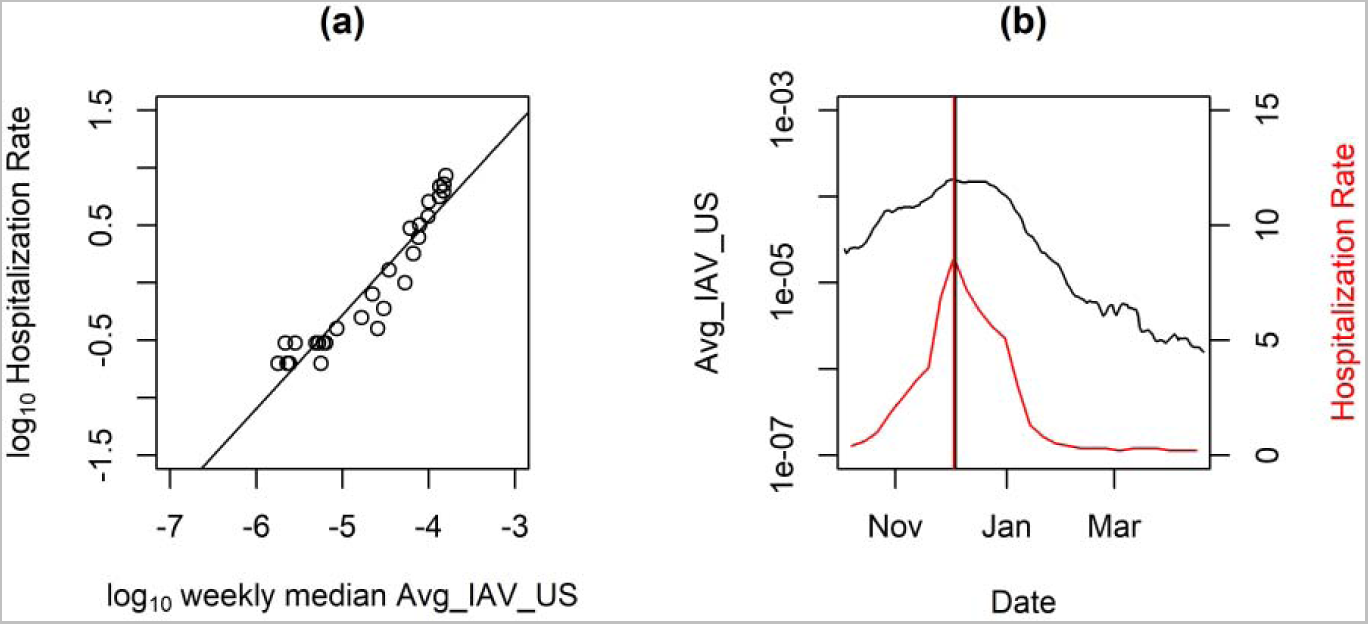
2022-2023 Overall FluServe-Net hospitalization rate (hospitalizations per 100,000 population) and Avg_IAV_US (a) linear relationship using log_10_ transformed values and weekly median Avg_IAV_US and (b) comparison of peak and intensity with vertical lines presenting time of peak.

#### 3.5.3 Linear relationship

We found a significant relationship between log_10_ Avg_IAV_State (expressed as the median of daily values each week) and log_10_ FluSurv-NET hospitalization rate (reported weekly) for each state (p < 0.001, R2 range of 0.74 to 0.90 across states), with a 0.61 to 0.80-unit increase in log_10_ hospitalization rate for every log_10_ increase in Avg_IAV_State (Figure S7 and Table S7). Considering the US aggregated inputs, we also found a significant relationship between log_10_ Avg_IAV_US (expressed as the median of daily values each week) and log_10_ overall hospitalization rate (reported weekly) (p < 0.001, R2 of 0.89), with a 0.81-unit increase in log_10_ overall hospitalization rate for every log_10_ increase in Avg_IAV_US (Figure 5a and Table S7).

## 4.0 Discussion

We used information on IAV and PMMoV RNA targets in municipal wastewater solids to estimate the 2022-2023 influenza season onset, offset, duration, peak and intensity. The CDC and other agencies use clinical data to estimate these characteristics.^6–8^ To compare the wastewater estimates to the clinical estimates, we aggregated the data from 163 WWTPs in 33 states to represent scales typically used for clinical data (e.g. state, HHS region, or national). Then, we estimated baseline concentrations of the aggregated wastewater data using approaches similar to those originally proposed for clinical data to identify onset, offset and duration, but with one year of data (or less) rather than multiple years.^25^

The wastewater baseline values were determined individually for HHS regions and states, as the ILI baseline values used by CDC are unique to each HHS region. The range of wastewater baseline values across the 5 regions covered approximately 1 log_10_ when set to correspond with ILI onset (minimum of 2.31×10^−6^ and maximum of 2.73×10^−5^). In comparison, the ranges of wastewater baseline values for HHS regions and states determined independently of ILI (baseline methods 2 - 4) covered less than 0.8 log_10_. The wastewater baseline values set to twice the minimum ranged between 5.70×10^−7^ and 3.46×10^−6^ across HHS regions and 8.1×10^−7^ and 3.54×10^−6^ across states. For reference, the difference between peak and baseline using twice the minimum for each state was 2 log_10_ or greater. Future work can explore if baseline values are applicable across states and/or spatial scales.

Regional results indicated that onset determined using wastewater and a baseline of the annual geometric mean was similar to the onset estimated using the ILI baseline; whereas, onset determined using wastewater and a baseline of the annual mean was later than ILI-estimated onsets. Using twice the minimum observation as wastewater baseline provided an early warning (between 30 and 75 days) compared to ILI-estimated onset. We applied the same approaches to estimate wastewater baseline values for states, for which ILI baseline data are not available. The pattern of onset across baseline methods mirrored the regional analysis; in addition, the baseline method influenced the estimated progression across states, in terms of the order of onset. Thus, for the 2022-2023 influenza season, the selection of baseline using wastewater data influenced the characterization (or interpretation) of onset in time and across space.

Characterizing the differences between baseline methods allows for public health officials to select a baseline that best matches their goals for identification of and communication about influenza season. For a wastewater baseline that aligns with expected increases in ILI, the geometric mean is best, as the onset of regions using the geometric mean of wastewater data aligned with the ILI-estimated onset. Alternatively, early warning of season onset may be desirable for public health decision making; in which case, a baseline based on the minimum observation may be useful. However, selecting the threshold using the minimum observation is challenging given the variation within each season (as illustrated by HHS R9 in Figure 1 for which summertime influenza activity interfered with identifying typical influenza season onset) and unknown variation between seasons (data not available to characterize).

Using information on the minimal observation is similar to the approach applied by Boehm et al.^13^ for identifying influenza wastewater events at individual wastewater treatment plants. In the present study, using the value of twice the minimal observation to identify IAV onset seemed to capture the entire periods of increasing and decreasing concentrations in wastewater during the seasonal IAV epidemic. However, in California, spatially aggregated wastewater levels remained above this baseline for the entire summer prior to the winter influenza season. California experienced higher than normal summertime influenza activity in 2022 after not having any influenza during the height of the COVID-19 pandemic^14^ which may have contributed to the high levels in wastewater in summer 2022. Choosing a slightly higher value (three times the minimum) as a baseline for California would have led to offset in September, followed closely by onset.

The definition of onset is also important from a practical perspective. We adopted the description of onset from CDC of the day when the measure was above the baseline value and remained there for at least two additional weeks. This definition precludes real-time determination of onset as it requires a two week period of data, which would retroactively identify an onset date at the beginning of the period when used in real time. Alternatively, a look-back window could be implemented, as applied by Boehm et al.^13^ for IAV, which provides a real-time estimation of an onset date assigned to the end of a two week period of data. Further work could investigate that definition.

We also compared the peak and intensity of the 2022-2023 influenza season using influenza hospitalization rates to that using wastewater data for states and the US. We found that the peaks estimated by wastewater data generally preceded the peaks determined by hospitalization rates for states by 6 to 14 days. The earlier peak is not surprising given that the wastewater data captures viral shedding from both asymptomatic and symptomatic infections, as well as from mild and severe infections whereas the hospitalization rate captures the most severe cases. The general agreement of peak combined with the strong relationship between Avg_IAV_State and hospitalization rate suggests one could use IAV RNA in wastewater solids as early warning for identifying peak hospitalization, especially for states with no clinical surveillance (e.g. data available for FL and TX for wastewater but not available in FluSurv-NET). The wastewater data could also be used at more localized scales to identify peak events, as has been illustrated previously at the building^11^, subsewershed^9^, and sewershed^13^ levels.

A strength of the wastewater data used in this analysis is wide coverage of WWTPs across the US. This allows analysis at a localized scale, as exemplified here by the state onset analysis and by onset analysis at the sewershed level elsewhere^13^. This analysis is enabled by the ability to aggregate information from individual WWTPs, which is possible due to the consistent methods applied to obtain IAV and PMMoV RNA concentrations across locations. Another strength is that the wastewater data captures IAV-specific mild illness and asymptomatic illness and is agnostic as to whether individuals seek medical care; whereas, the ILI data captures data from individuals with various respiratory illnesses seeking medical care, which resulted in the exclusion of the ILI data collected over the pandemic in the calculation of the baseline.^15^ This is particularly important as COVID-19 surveillance is expected to transition to a model that is more similar to influenza surveillance, and there is a need to distinguish IAV and COVID-19 outbreaks that share similar ailments and symptoms.

Limitations of using wastewater data to characterize influenza season include that wastewater data cannot be disaggregated by age of population, vaccine status of population, or severity of disease. WWTPS participation is voluntary; therefore, coverage may vary over time and sample sites are not based on a strategic sampling plan to provide optimal coverage across the US, regions, or states. Additionally, the wastewater data lacks the historical record that clinical data provide.

Wastewater settled solids data represent an additional surveillance indicator of influenza that can be used to characterize the influenza season along with clinical data across scales currently unavailable using clinical data (e.g., US down to sewershed). Whereas the rates of influenza-associated hospitalizations are a product of the transmissibility and the clinical severity of influenza,^7^ wastewater data may be considered as a proxy to transmissibility, similar to ILI, but IAV specific. As additional data are collected over multiple influenza seasons, the approaches introduced in this work can be refined to better capture the variation in IAV and PMMoV targets in municipal wastewater solids across seasons and further analysis can be conducted such as the moving epidemic method or aggregate average method^25^ to evaluate IAV onset and severity.^7^

## Data Availability

Wastewater data are publicly available at the Stanford Digital Repository: https://doi.org/10.25740/mf441vr6745.

## Author Contributions

The manuscript was written through contributions of all authors. All authors have given approval to the final version of the manuscript. MS and ALB computation; MS and ABB writing; MS, ALB, MW, and ABB contributed to the conception and editing of this article.

## ACKNOWLEDGMENT

We acknowledge the numerous people who contributed to wastewater sample collection.

## Notes

### Competing Interest Statement

The authors have declared no competing interest.

### Funding Statement

This work was supported by gifts from the CDC Foundation and the Sergey Brin Family Foundation to ABB.

### Author Declarations

The aggregated hospitalization and syndromic data were opening available before the initiation of this study. They can be access using the following links: Laboratory-Confirmed Influenza Hospitalizations (https://gis.cdc.gov/GRASP/Fluview/FluHospRates.html) and National, Regional, and State Level Outpatient Illness and Viral Surveillance (https://gis.cdc.gov/grasp/fluview/fluportaldashboard.html).

