## Supporting information for "United States influenza 2022-2023 season characteristics as inferred from wastewater solids, influenza hospitalization and syndromic data"

### Methods

**Additional details on pre-analytical methods.** The pre-analytical methods have been registered at protocols.io[1] and described in other publications[2]. Pre-analytical processing occurred as soon as the samples were received at the laboratory. In brief, wastewater solids are dewatered using centrifugation. Thereafter, one aliquot is taken to measure the dry weight using an oven. Another aliquot is added to bovine-coronavirus-spiked DNA/RNA shield (Zymo, Irvine CA) so the final concentration is ~75 mg/ml; at this concentration, we observe minimal inhibition of the assays.[2] The solution is then homogenized using grinding balls, and then centrifuged again. The nucleic-acids are then extracted from an aliquot of the supernatant using a commercial extraction kit. This is done in 6 or 10 replicates so that there are 6 or 10 distinct nucleic-acid extractions from each solids sample. The only sites that used 10 replicates are Gilroy, CA; Oceanside San Francisco, CA; Palo Alto, CA; Redwood City, CA; Sacramento, CA; San Jose, CA; Southeast San Francisco, CA; and Sunnyvale, CA. The rest of the WWTPs used 6 replicates.

**Additional details on the analytical methods.** Nucleic-acids were processed immediately with no storage. We measured concentrations of the influenza M1 gene using a primer and probe set that has been previously used in wastewater[2] and originally published by the CDC[3] using droplet digital RT-PCR. The nucleic-acid extracts were used as template neat (not diluted). Each nucleic-acid extract was run in its own well so that 6 or 10 replicate wells were run per sample. The IAV assay was multiplexed with other assays, and those assays changed over the period of the project as public health needs changed and new research to support the monitoring of additional disease targets became available; the assays IAV was multiplexed with are provided in Figure S1. PMMoV and BCoV were assayed using the same methods as previously described[2]; nucleic-acids were diluted 1:100 as template; two or 10 wells were run per sample; the sites where 10 wells were run are the same as those mentioned in the previous section. The specific methods have been described in detail elsewhere[2]. The replicate wells were merged for analysis and the methods for thresholding were described by Boehm et al.[2].

**QA/QC.** Extraction positive and negative controls, as well as RT-PCR positive and negative controls were included on each plate. All positive and negative controls were positive and negative, respectively. More information on the QA/QC are provided by Boehm et al.[2] regarding the preparation of the controls. In order for a sample to be counted as positive, it had to have 3 positive droplets across the merged wells. This is equivalent to a concentration of about 500-1000 copies per gram dry weight; the range reflects the varied dry weights of the solids, and whether 6 or 10 wells were used for the sample.

**Linear relationship.** Linear relationships were evaluated between $\log_{10}$ transformed a) weekly median Avg_IAV_State and hospitalization rate at the state level and b) weekly median Avg_IAV_US and overall hospitalization rate for the CDC influenza surveillance period beginning 10/2/2022 but shortened to 4/15/2023, the point in time when states reached offset (see manuscript results). Hospitalization rates of 0.0 hospitalizations per 100,000 population were set to 0.05 for transformation. Weekly median Avg_IAV_State below the LOD were removed (CA = 0, CO = 5, GA = 2, MN = 4, MI= 3, and UT = 2 between 10/2/2022 and 4/15/2023).

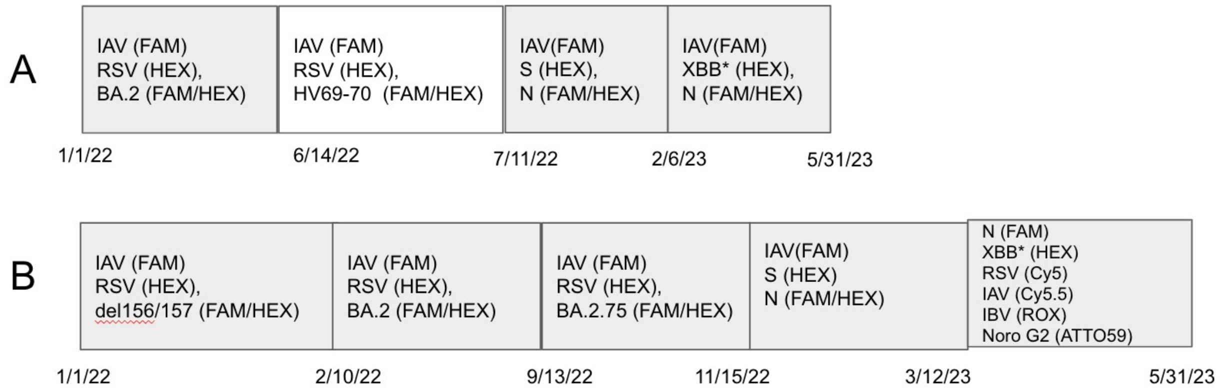

**Figure S1.** A schematic of how assays were multiplexed during different periods of the project.

Row A is for all WWTPS in the study except for Gilroy, CA; Oceanside San Francisco, CA; Palo Alto, CA; Redwood City, CA; Sacramento, CA; San Jose, CA; Southeast San Francisco, CA; and Sunnyvale, CA. Row B is for Gilroy, CA; Oceanside San Francisco, CA; Palo Alto, CA; Redwood City, CA; Sacramento, CA; San Jose, CA; Southeast San Francisco, CA; and Sunnyvale, CA. The date at the bottom left side of each box is the start date and the date near the right edge of the box is the approximate end date (based on the date the assay was stopped in the lab, the date associated with the last sample run could be different depending on the date it was processed in the lab). All probes contained fluorescent molecules as indicated in parentheses: FAM, 6-fluorescein amidite; HEX, hexachloro-fluorescein, Cy5, Cyanine-5; Cy5.5, Cyanine5.5; ROX, carboxyrhodamine; and ATTO59. If the box is white, the annealing temperature is 59 °C and if gray, it is 61 °C. N and S are assays targeting those genes in SARS-CoV-2. HV69-70 is the assay targeting the deletion 69/70 in the S gene characteristic of Alpha, and various Omicron variants. BA.2.75 is the assay targeting the adjunct SNPs in the S gene characteristic of BA.2.75. RSV is the assay targeting a gene in the RSV genome. BA.2 is the assay targeting the set of deletions LPPA24S characteristic of BA.2. IBV is the assay targeting influenza B. XBB* is an assay targeted adjacent SNPs in the SARS-CoV-2 XBB* sublineages. Noro G2 is an assay targeting the genome of human norovirus GII.

(1) Topol, A.; Wolfe, M.; White, B.; Wigginton, K.; Boehm, A. High Throughput Pre-Analytical Processing of Wastewater Settled Solids for SARS-CoV-2 RNA Analyses. *protocols.io* **2021**.
(2) Boehm, A. B.; Wolfe, M. K.; Wigginton, K. R.; Bidwell, A.; White, B. J.; Hughes, B.; Duong, D.; Chan-Herur, V.; Bischel, H. N.; Naughton, C. C. Human Viral Nucleic Acids Concentrations in Wastewater Solids from Central and Coastal California USA. *Scientific Data* **2023**, *10* (1), 396.
(3) CDC. *Labs*. Centers for Disease Control and Prevention. https://www.cdc.gov/coronavirus/2019-ncov/lab/multiplex.html (accessed 2023-09-08).

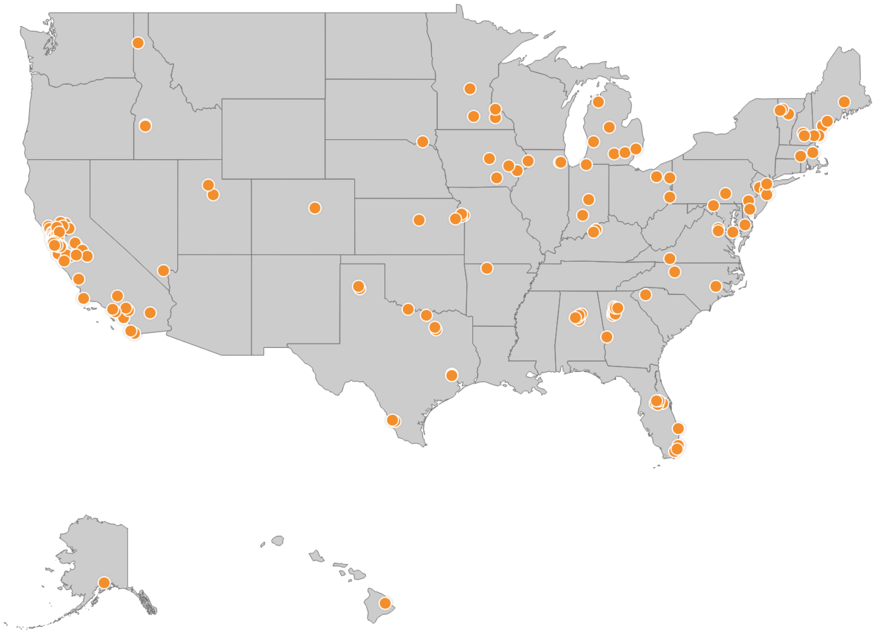

**Figure S2**. Map of 163 WWTPs. Each orange dot indicates a WWTP location.

**Table S1.** Details of the 163 WWTPs included in this study.

The state where the WWTP is located, additional details regarding HHS region, plant location, site name, sample type, reported population served, and sample IAV descriptive statistics. ND: Non-detect, Min: Minimum, Max: Maximum, Med: Median. If the sample type is listed as "Solids" then that means the site provided samples of settled solids from the primary clarifier. If the sample type is labeled as "Solids (Settling w/ Imhoff cones on site)" the solids were obtained from raw influent using an Imhoff cone on site. If the sample type is labeled "Liquid", then the solids were obtained in the laboratory by allowing the raw influent to settle for 10-15 mins, and using a serological pipette to aspirate the settled solids into a falcon tube.

| State | HHS Region | Plant | Site Name | Sample Type | Population Served | Sample Count | Sample Start | Sample End | Min IAV (cp/g) | Max IAV (cp/g) | Med IAV (cp/g) |
|---|---|---|---|---|---|---|---|---|---|---|---|
| Alabama | HHS4 | Bessemer, AL | Valley Creek Water Reclamation Facility | Liquids | 225,000 | 125 | 8/15/22 | 5/31/23 | ND | 289,429 | 0 |
| | | Cahaba River, Birmingham, AL | Cahaba River Water Reclamation Facility | Liquids | 95,000 | 107 | 8/15/22 | 5/29/23 | ND | 542,279 | 3,441 |
| | | Fultondale, AL | Five Mile Creek Water Reclamation Facility | Liquids | 77,000 | 95 | 8/22/22 | 5/31/23 | ND | 112,939 | 0 |
| | | Pinson, AL | Turkey Creek Water Reclamation Facility | Liquids | 30,000 | 93 | 8/15/22 | 5/31/23 | ND | 426,025 | 0 |
| | | Village Creek, Birmingham, AL | Village Creek Water Reclamation Facility | Liquids | 200,000 | 121 | 8/16/23 | 5/30/23 | ND | 112,887 | 0 |

| State | HHS Region | Plant | Site Name | Sample Type | Population Served | Sample Count | Sample Start | Sample End | Min IAV (cp/g) | Max IAV (cp/g) | Med IAV (cp/g) |
|---|---|---|---|---|---|---|---|---|---|---|---|
| Alaska | HHS10 | Anchorage, AK | John M. Asplund Water Pollution Control Facility | Liquids | 220,000 | 4 | 5/24/23 | 5/31/23 | ND | 26,135 | 5,649 |
| Arkansas | HHS6 | Harrison, AR | City of Harrison Wastewater Treatment Plant | Liquids | 15,000 | 15 | 4/23/23 | 5/31/23 | ND | 0 | 0 |
| California | HHS9 | Coastal, Laguna Niguel, CA | Coastal Treatment Plant | Liquids | 48,000 | 69 | 12/21/22 | 5/30/23 | ND | 55,602 | 0 |
| | | CODIGA, Stanford, CA | CODIGA | Solids (Settling w/ Imhoff cones on site) | 10,000 | 396 | 1/5/22 | 5/31/23 | ND | 126,943 | 0 |
| | | Contra Costa County, CA | Central Contra Costa Sanitary District | Solids | 484,800 | 171 | 3/21/22 | 5/30/23 | ND | 197,261 | 2,552 |
| | | Davis, CA | City of Davis Wastewater Treatment Plant | Solids | 68,000 | 426 | 1/5/22 | 5/31/23 | ND | 105,010 | 1,169 |
| | | Esparto, CA | Esparto Wastewater | Liquids | 4,006 | 72 | 12/2/22 | 5/30/23 | ND | 90,468 | 0 |

| State | HHS Region | Plant | Site Name | Sample Type | Population Served | Sample Count | Sample Start | Sample End | Min IAV (cp/g) | Max IAV (cp/g) | Med IAV (cp/g) |
|---|---|---|---|---|---|---|---|---|---|---|---|
| | | | Treatment Facility | | | | | | | | |
| | | Fairfield, CA | Fairfield-Suisun Sewer District | Solids | 155,000 | 105 | 9/28/22 | 5/31/23 | ND | 86,695 | 2,714 |
| | | Fremont, CA | [Fremont Basin] - Raymond A. Boege Alvarado WWTP | Liquids | 229,476 | 56 | 11/1/22 | 5/30/23 | ND | 178,713 | 0 |
| | | Gilroy, CA | South County Regional Wastewater Authority | Solids (Settling w/ Imhoff cones on site) | 110,338 | 511 | 1/5/22 | 5/31/23 | ND | 264,912 | 0 |
| | | Half Moon Bay, CA | Sewer Authority Mid-Coastside | Liquids | 28,000 | 150 | 4/27/22 | 5/31/23 | ND | 210,648 | 0 |
| | | Hollister, CA | City of Hollister Domestic Water Recycling Facility | Liquids | 42,000 | 106 | 9/14/22 | 5/31/23 | ND | 1,504,415 | 0 |
| | | Indio, CA | Valley Sanitary District | Solids | 91,765 | 121 | 8/24/22 | 5/31/23 | ND | 155,979 | 0 |

| State | HHS Region | Plant | Site Name | Sample Type | Population Served | Sample Count | Sample Start | Sample End | Min IAV (cp/g) | Max IAV (cp/g) | Med IAV (cp/g) |
|---|---|---|---|---|---|---|---|---|---|---|---|
| | | JB Latham, Laguna Niguel, CA | JB Latham Treatment Plant | Liquids | 120,000 | 69 | 12/23/22 | 5/30/23 | ND | 292,870 | 0 |
| | | Lancaster, CA | Lancaster Water Reclamation Plant | Liquids | 200,000 | 112 | 9/12/22 | 5/30/23 | ND | 301,712 | 0 |
| | | Las Gallinas, San Rafael, CA | Las Gallinas Valley Sanitary District | Liquids | 30,000 | 129 | 8/8/22 | 5/31/23 | ND | 217,611 | 0 |
| | | Lompoc, CA | Lompoc Regional Wastewater Reclamation Plant | Liquids | 69,290 | 131 | 8/1/22 | 5/31/23 | ND | 395,660 | 0 |
| | | Los Angeles County, CA | Joint Water Pollution Control Plant | Liquids | 3,500,000 | 216 | 1/9/22 | 5/31/23 | ND | 187,289 | 0 |
| | | Los Angeles, CA | Hyperion Water Reclamation Plant (HWRP) | Liquids | 4,000,000 | 119 | 8/28/22 | 5/31/23 | ND | 163,912 | 0 |

| State | HHS Region | Plant | Site Name | Sample Type | Population Served | Sample Count | Sample Start | Sample End | Min IAV (cp/g) | Max IAV (cp/g) | Med IAV (cp/g) |
|---|---|---|---|---|---|---|---|---|---|---|---|
| | | Los Banos, CA | Los Banos Wastewater Treatment Plant | Liquids | 42,000 | 79 | 12/2/22 | 5/30/23 | ND | 212,297 | 0 |
| | | Madera, CA | City of Madera, Wastewater Treatment Plant | Solids | 67,944 | 35 | 3/6/23 | 5/31/23 | ND | 2,054 | 0 |
| | | Marina, CA | Monterey One Water - Regional Treatment Plant | Liquids | 262,000 | 74 | 11/27/22 | 5/30/23 | ND | 205,231 | 0 |
| | | Merced, CA | Merced Wastewater Treatment Plant | Solids (Settling w/ Imhoff cones on site) | 91,000 | 193 | 1/5/22 | 5/31/23 | ND | 96,641 | 0 |
| | | Mill Valley, CA | Sewerage Agency of Southern Marin Wastewater Treatment Plant | Solids | 30,000 | 73 | 12/12/22 | 5/30/23 | ND | 98,803 | 2,507 |
| | | Modesto, CA | Modesto's Sutter Primary | Solids | 230,000 | 188 | 1/5/22 | 5/31/23 | ND | 103,433 | 0 |

| State | HHS Region | Plant | Site Name | Sample Type | Population Served | Sample Count | Sample Start | Sample End | Min IAV (cp/g) | Max IAV (cp/g) | Med IAV (cp/g) |
|---|---|---|---|---|---|---|---|---|---|---|---|
| | | | Treatment Facility | | | | | | | | |
| | | Napa, CA | Soscol Water Recycling Facility | Solids | 83,300 | 103 | 9/26/22 | 5/31/23 | ND | 223,313 | 2,549 |
| | | Newark, CA | [Newark Basin] - Raymond A. Boege Alvarado WWTP | Liquids | 47,229 | 57 | 11/3/22 | 5/30/23 | ND | 386,425 | 0 |
| | | Novato, CA | Novato Sanitary District | Liquids | 53,000 | 145 | 6/20/22 | 5/31/23 | ND | 1,842,329 | 0 |
| | | Oakland, CA | East Bay Municipal Utility District | Solids | 740,000 | 204 | 2/15/22 | 5/31/23 | ND | 170,822 | 298 |
| | | Oceanside, San Francisco, CA | Oceanside Water Pollution Control Plant | Solids | 250,000 | 492 | 1/5/22 | 5/31/23 | ND | 161,365 | 1,369 |
| | | Ontario, CA | Regional Water Recycling Plant No.1 (RP-1) | Solids | 890,000 | 170 | 4/25/22 | 5/31/23 | ND | 255,134 | 0 |

| State | HHS Region | Plant | Site Name | Sample Type | Population Served | Sample Count | Sample Start | Sample End | Min IAV (cp/g) | Max IAV (cp/g) | Med IAV (cp/g) |
|---|---|---|---|---|---|---|---|---|---|---|---|
| | | Pacifica, CA | Calera Creek Water Recycling Plant | Liquids | 40,000 | 94 | 10/17/22 | 5/31/23 | ND | 1,199,912 | 0 |
| | | Palo Alto, CA | Palo Alto Regional Water Quality Control Plant | Solids | 236,000 | 511 | 1/5/22 | 5/31/23 | ND | 15,438,760 | 4,096 |
| | | Paso Robles, CA | City of Paso Robles Wastewater Treatment Plant | Solids | 31,037 | 215 | 1/11/22 | 5/30/23 | ND | 144,647 | 1,636 |
| | | Petaluma, CA | Ellis Creek Water Recycling Facility | Liquids | 65,000 | 141 | 6/28/22 | 5/30/23 | ND | 471,071 | 0 |
| | | Redwood City, CA | Silicon Valley Clean Water | Solids | 199,000 | 512 | 1/5/22 | 5/31/23 | ND | 106,645 | 1,531 |
| | | Regional, Laguna Niguel, CA | Regional Treatment Plant | Liquids | 129,000 | 68 | 12/21/22 | 5/30/23 | ND | 97,703 | 0 |

| State | HHS Region | Plant | Site Name | Sample Type | Population Served | Sample Count | Sample Start | Sample End | Min IAV (cp/g) | Max IAV (cp/g) | Med IAV (cp/g) |
|---|---|---|---|---|---|---|---|---|---|---|---|
| | | Riverside, CA | Riverside Water Quality Control Plant | Liquids | 350,000 | 55 | 1/25/23 | 5/31/23 | ND | 1,027,468 | 0 |
| | | Sacramento, CA | Sacramento Regional Wastewater Treatment Plant | Solids | 1,480,000 | 512 | 1/5/22 | 5/31/23 | ND | 107,875 | 1,601 |
| | | San Diego, CA | E.W. Blom Point Loma Wastewater Treatment Plant | Liquids | 2,200,000 | 125 | 8/10/22 | 5/31/23 | ND | 186,150 | 0 |
| | | San Jose, CA | San Jose-Santa Clara Regional Wastewater Facility | Solids | 1,500,000 | 512 | 1/5/22 | 5/31/23 | ND | 167,638 | 2,786 |
| | | San Leandro, CA | City of San Leandro Water Pollution Control Plant | Liquids | 50,000 | 107 | 9/27/22 | 5/31/23 | ND | 102,259 | 0 |
| | | San Mateo, CA | City of San Mateo & Estero M.I.D. Water | Solids | 150,000 | 129 | 7/6/22 | 5/31/23 | ND | 108,089 | 0 |

| State | HHS Region | Plant | Site Name | Sample Type | Population Served | Sample Count | Sample Start | Sample End | Min IAV (cp/g) | Max IAV (cp/g) | Med IAV (cp/g) |
|---|---|---|---|---|---|---|---|---|---|---|---|
| | | | Quality Control Plant | | | | | | | | |
| | | San Rafael, CA | Central Marin Sanitation Agency | Liquids | 104,250 | 73 | 8/22/22 | 5/30/23 | ND | 219,843 | 6,258 |
| | | Santa Cruz County, CA | City of Santa Cruz WTF - County Influent | Solids (Settling w/ Imhoff cones on site) | 160,000 | 177 | 4/3/22 | 5/30/23 | ND | 148,269 | 0 |
| | | Santa Cruz, CA | City of Santa Cruz WTF - City Influent | Solids (Settling w/ Imhoff cones on site) | 160,000 | 176 | 4/3/22 | 5/30/23 | ND | 507,690 | 0 |
| | | Santa Rosa, CA | City of Santa Rosa, Laguna Treatment Plant | Solids (Settling w/ Imhoff cones on site) | 230,000 | 124 | 8/11/22 | 5/31/23 | ND | 125,061 | 0 |

| State | HHS Region | Plant | Site Name | Sample Type | Population Served | Sample Count | Sample Start | Sample End | Min IAV (cp/g) | Max IAV (cp/g) | Med IAV (cp/g) |
|---|---|---|---|---|---|---|---|---|---|---|---|
| | | Sausalito, CA | Sausalito-Marin City Sanitary District | Liquids | 18,000 | 104 | 8/8/22 | 5/23/23 | ND | 357,317 | 0 |
| | | South San Diego, CA | South Bay International Wastewater Treatment Plant | Solids | 1,600,000 | 36 | 12/14/22 | 3/10/23 | ND | 14,871 | 1,819 |
| | | Southeast San Francisco, CA | Southeast San Francisco | Solids | 750,000 | 370 | 5/20/22 | 5/31/23 | ND | 102,331 | 2,363 |
| | | Sunnyvale, CA | City of Sunnyvale Water Pollution Control Plant | Solids | 153,000 | 503 | 1/5/22 | 5/31/23 | ND | 107,816 | 2,153 |
| | | Turlock, CA | Turlock Regional Water Quality Control Facility | Liquids | 86,000 | 78 | 12/2/22 | 5/31/23 | ND | 63,168 | 0 |
| | | Union City, CA | [Union City Basin] - Raymond A. Boege Alvarado WWTP | Liquids | 68,150 | 57 | 11/1/22 | 5/30/23 | ND | 107,183 | 0 |

| State | HHS Region | Plant | Site Name | Sample Type | Population Served | Sample Count | Sample Start | Sample End | Min IAV (cp/g) | Max IAV (cp/g) | Med IAV (cp/g) |
|---|---|---|---|---|---|---|---|---|---|---|---|
| | | University of California, Davis, CA | UC Davis | Solids | n/a | 378 | 1/5/22 | 5/30/23 | ND | 83,471 | 0 |
| | | Vallejo, CA | Vallejo Flood and Wastewater District Wastewater Treatment Plant | Liquids | 121,000 | 108 | 9/20/22 | 5/31/23 | ND | 266,271 | 946 |
| | | West Contra Costa County, CA | West County Wastewater District | Liquids | 100,000 | 128 | 5/12/22 | 5/30/23 | ND | 390,836 | 2,708 |
| | | West Railroad, San Rafael, CA | Central Marin Sanitation Agency - West Railroad | Liquids | 25,000 | 75 | 8/24/22 | 5/30/23 | ND | 238,377 | 0 |
| | | Windsor, CA | Windsor Wastewater Treatment, Reclamation, and Disposal Facility | Liquids | 28,000 | 63 | 12/16/22 | 5/31/23 | ND | 33,751 | 0 |

| State | HHS Region | Plant | Site Name | Sample Type | Population Served | Sample Count | Sample Start | Sample End | Min IAV (cp/g) | Max IAV (cp/g) | Med IAV (cp/g) |
|---|---|---|---|---|---|---|---|---|---|---|---|
| | | Winters, CA | Winters - East Street Pump Station | Liquids | 7,286 | 72 | 12/2/22 | 5/31/23 | ND | 130,150 | 0 |
| | | Woodland, CA | Woodland Water Pollution Control Facility | Liquids | 59,000 | 78 | 12/2/22 | 5/31/23 | ND | 112,381 | 0 |
| Colorado | HHS8 | North, Parker, CO | Parker Water and Sanitation District North Water Reclamation Facility | Liquids | 35,000 | 159 | 5/16/22 | 5/31/23 | ND | 162,484 | 0 |
| | | South, Parker, CO | Parker Water and Sanitation District South Water Reclamation Facility | Liquids | 25,000 | 160 | 5/16/22 | 5/31/23 | ND | 182,314 | 0 |
| Delaware | HHS3 | Seaford, DE | Seaford Wastewater Treatment Facility | Solids | 13,172 | 48 | 2/8/23 | 5/31/23 | ND | 3,807 | 0 |

| State | HHS Region | Plant | Site Name | Sample Type | Population Served | Sample Count | Sample Start | Sample End | Min IAV (cp/g) | Max IAV (cp/g) | Med IAV (cp/g) |
|---|---|---|---|---|---|---|---|---|---|---|---|
| Florida | HHS4 | Altamonte Springs, FL | Altamonte Springs Regional Water Reclamation Facility | Liquids | 95,000 | 84 | 10/24/22 | 5/31/23 | ND | 575,996 | 3,875 |
| | | Eastern, Orange County, FL | Eastern Water Reclamation Facility | Solids (Settling w/ Imhoff cones onsite) | 195,299 | 168 | 4/3/22 | 5/31/23 | ND | 42,802 | 0 |
| | | Jupiter, FL | Loxahatchee River Environmental Control District | Liquids | 90,000 | 109 | 9/14/22 | 5/31/23 | ND | 59,598 | 0 |
| | | Key Biscayne, FL | MDWASD Central District WWTP | Liquids | 829,725 | 51 | 1/22/23 | 5/31/23 | ND | 11,087 | 0 |
| | | North Miami, FL | MDWASD North District WWTF | Liquids | 776,150 | 58 | 1/16/23 | 5/31/23 | ND | 5,556 | 0 |
| | | Northwest, Orange County, FL | Northwest Water Reclamation Facility | Solids (Settling w/ Imhoff | 66,690 | 169 | 4/3/22 | 5/31/23 | ND | 59,678 | 0 |

| State | HHS Region | Plant | Site Name | Sample Type | Population Served | Sample Count | Sample Start | Sample End | Min IAV (cp/g) | Max IAV (cp/g) | Med IAV (cp/g) |
|---|---|---|---|---|---|---|---|---|---|---|---|
| | | | | cones onsite) | | | | | | | |
| | | South Miami, FL | MDWASD South District WWTF | Liquids | 920,528 | 55 | 1/15/23 | 5/31/23 | ND | 32,313 | 0 |
| | | South, Orange County, FL | South Water Reclamation Facility | Solids (Settling w/ Imhoff cones onsite) | 183,009 | 168 | 4/3/22 | 5/31/23 | ND | 83,923 | 0 |
| | | Southwest, Orange County, FL | Hamlin Water Reclamation Facility | Liquids | 50,000 | 79 | 10/27/22 | 5/30/23 | ND | 130,356 | 0 |
| Georgia | HHS4 | Big Creek, Roswell, GA | Big Creek Water Reclamation Facility | Liquids | 189,593 | 144 | 6/26/22 | 5/31/23 | ND | 142,953 | 985 |
| | | College Park, GA | Camp Creek Water Reclamation Facility | Liquids | 73,821 | 143 | 6/26/22 | 5/31/23 | ND | 95,002 | 2,847 |

| State | HHS Region | Plant | Site Name | Sample Type | Population Served | Sample Count | Sample Start | Sample End | Min IAV (cp/g) | Max IAV (cp/g) | Med IAV (cp/g) |
|---|---|---|---|---|---|---|---|---|---|---|---|
| | | Columbus, GA | South Columbus Water Resources Facility | Solids | 278,000 | 100 | 8/15/22 | 5/24/23 | ND | 92,572 | 4,171 |
| | | Johns Creek, Roswell, GA | Johns Creek Environmental Campus | Liquids | 84,486 | 143 | 6/26/22 | 5/31/23 | ND | 169,404 | 4,213 |
| | | Little River, Roswell, GA | Little River Water Reclamation Facility | Liquids | 12,818 | 144 | 6/27/22 | 5/31/23 | ND | 294,380 | 0 |
| | | RM Clayton, Atlanta, GA | RM Clayton Water Reclamation Center | Liquids | 294,660 | 86 | 10/30/22 | 5/30/23 | ND | 56,590 | 1,589 |
| | | South River, Atlanta, GA | South River Water Reclamation Center | Liquids | 105,160 | 85 | 10/30/22 | 5/30/23 | ND | 240,789 | 3,553 |
| | | Utoy Creek, Atlanta, GA | Utoy Creek Water | Liquids | 70,887 | 85 | 10/30/22 | 5/30/23 | ND | 136,184 | 0 |

| State | HHS Region | Plant | Site Name | Sample Type | Population Served | Sample Count | Sample Start | Sample End | Min IAV (cp/g) | Max IAV (cp/g) | Med IAV (cp/g) |
|---|---|---|---|---|---|---|---|---|---|---|---|
| | | | Reclamation Center | | | | | | | | |
| Hawaii | HHS9 | Hilo, HI | Hilo Wastewater Treatment Plant | Liquids | 16,257 | 11 | 5/8/23 | 5/31/23 | ND | 5,615 | 0 |
| Idaho | HHS10 | Coeur d'Alene, ID | City of Coeur d'Alene Water Resource Recovery Facility | Solids | 50,540 | 197 | 1/21/22 | 5/30/23 | ND | 210,676 | 0 |
| | | Lander Street, Boise, ID | Lander Street Water Renewal Facility | Liquids | 108,556 | 59 | 1/16/23 | 5/31/23 | ND | 36,882 | 0 |
| | | West Boise, ID | West Boise Water Renewal Facility | Liquids | 186,901 | 59 | 1/16/23 | 5/31/23 | ND | 38,293 | 0 |
| Illinois | HHS5 | Glen Ellyn, IL | Glenbard Wastewater Authority | Solids (Settling w/ Imhoff cones onsite) | 86,000 | 117 | 8/4/22 | 5/30/23 | ND | 115,106 | 0 |

| State | HHS Region | Plant | Site Name | Sample Type | Population Served | Sample Count | Sample Start | Sample End | Min IAV (cp/g) | Max IAV (cp/g) | Med IAV (cp/g) |
|---|---|---|---|---|---|---|---|---|---|---|---|
| | | Wheaton, IL | Wheaton Sanitary District | Solids (Settling w/ Imhoff cones onsite) | 63,000 | 108 | 9/14/22 | 5/31/23 | ND | 95,105 | 3,524 |
| Indiana | HHS5 | Bloomington, IN | Dillman Road WWTP | Liquids | 56,090 | 113 | 8/15/22 | 5/31/23 | ND | 237,971 | 0 |
| | | Carmel, IN | City of Carmel WWTP | Solids | 86,000 | 14 | 5/1/23 | 5/31/23 | ND | 0 | 0 |
| | | Downtown, Jeffersonville, IN | Jeffersonville Downtown WWTP | Liquids | 25,000 | 89 | 10/26/22 | 5/31/23 | ND | 388,202 | 0 |
| | | North, Jeffersonville, IN | North Water Reclamation Facility | Liquids | 25,000 | 87 | 10/26/22 | 5/31/23 | ND | 226,454 | 0 |
| | | South Bend, IN | City of South Bend Wastewater Treatment Plant | Liquids | 130,000 | 98 | 9/11/22 | 5/30/23 | ND | 314,228 | 0 |
| Iowa | HHS7 | Clinton, IA | City of Clinton | Solids | 29,300 | 58 | 1/18/23 | 5/31/23 | ND | 9,988 | 0 |

| State | HHS Region | Plant | Site Name | Sample Type | Population Served | Sample Count | Sample Start | Sample End | Min IAV (cp/g) | Max IAV (cp/g) | Med IAV (cp/g) |
|---|---|---|---|---|---|---|---|---|---|---|---|
| | | Coralville, IA | Coralville Wastewater Treatment Facility | Liquids | 23,000 | 53 | 1/23/23 | 5/31/23 | ND | 4,375,144 | 0 |
| | | Marshalltown, IA | City of Marshalltown Water Pollution Control Plant | Liquids | 27,400 | 55 | 1/22/23 | 5/30/23 | ND | 403,633 | 9,514 |
| | | Muscatine, IA | Muscatine STP | Solids | 24,400 | 73 | 12/12/22 | 5/31/23 | ND | 154,346 | 0 |
| | | Ottumwa, IA | Ottumwa WPCF | Liquids | 25,529 | 72 | 12/16/22 | 5/30/23 | ND | 218,435 | 14,008 |
| Kansas | HHS7 | Kaw Point, Kansas City, KS | Municipal Wastewater Treatment Plant No. 1 (Kaw Point) | Solids (Settling w/ Imhoff cones onsite) | 90,000 | 60 | 1/10/23 | 5/31/23 | ND | 11,159 | 0 |
| | | Lawrence, KS | Lawrence Kansas River Wastewater Treatment Facility | Solids (Settling w/ Imhoff cones onsite) | 80,000 | 121 | 8/1/22 | 5/31/23 | ND | 167,592 | 0 |

| State | HHS Region | Plant | Site Name | Sample Type | Population Served | Sample Count | Sample Start | Sample End | Min IAV (cp/g) | Max IAV (cp/g) | Med IAV (cp/g) |
|---|---|---|---|---|---|---|---|---|---|---|---|
| | | P20, Kansas City, KS | Kansas City Treatment Plant #20 | Solids (Settling w/ Imhoff cones onsite) | 35,000 | 61 | 1/10/23 | 5/31/23 | ND | 11,238 | 0 |
| | | Salina, KS | Salina Wastewater Treatment Plant | Solids | 47,000 | 128 | 8/8/22 | 5/31/23 | ND | 232,574 | 0 |
| | | Wolcott, Kansas City, KS | Wolcott Wastewater Treatment Facility | Liquids | 15,000 | 59 | 1/9/23 | 5/31/23 | ND | 60,947 | 0 |
| Kentucky | HHS4 | Louisville, KY | Morris Forman Water Quality Treatment Center | Solids (Settling w/ Imhoff cones onsite) | 423,913 | 64 | 2/24/22 | 5/25/23 | ND | 92,926 | 6,996 |
| Maine | HHS1 | Bangor, ME | City of Bangor Wastewater Treatment Plant | Liquids | 40,000 | 7 | 5/15/23 | 5/31/23 | ND | 13,063 | 0 |
| | | Brunswick, ME | Brunswick Sewer District | Liquids | 10,000 | 68 | 11/29/22 | 5/31/23 | ND | 1,387,797 | 0 |

| State | HHS Region | Plant | Site Name | Sample Type | Population Served | Sample Count | Sample Start | Sample End | Min IAV (cp/g) | Max IAV (cp/g) | Med IAV (cp/g) |
|---|---|---|---|---|---|---|---|---|---|---|---|
| | | Portland, ME | Portland Water District (East End Wastewater Treatment Facility) | Liquids | 65,000 | 89 | 9/2/22 | 5/31/23 | ND | 333,422 | 5,935 |
| | | York, ME | York Sewer District | Liquids | 10,000 | 55 | 1/18/23 | 5/31/23 | ND | 27,722 | 0 |
| Maryland | HHS3 | Hagerstown, MD | Hagerstown Wastewater Treatment Plant | Liquids | 90,000 | 68 | 12/14/22 | 5/31/23 | ND | 109,922 | 0 |
| | | Hollywood, MD | Marlay Taylor Water Reclamation Facility | Liquids | 55,000 | 54 | 1/4/23 | 5/31/23 | ND | 11,018 | 0 |
| Massachusetts | HHS1 | Boston, MA | Deer Island Treatment Plant | Solids | 2,400,000 | 70 | 12/12/22 | 5/31/23 | ND | 91,150 | 2,616 |
| | | Millbury, MA | Upper Blackstone Clean Water | Liquids | 250,000 | 39 | 2/27/23 | 5/30/23 | ND | 13,995 | 0 |

| State | HHS Region | Plant | Site Name | Sample Type | Population Served | Sample Count | Sample Start | Sample End | Min IAV (cp/g) | Max IAV (cp/g) | Med IAV (cp/g) |
|---|---|---|---|---|---|---|---|---|---|---|---|
| Michigan | HHS5 | Ann Arbor, MI | City of Ann Arbor Wastewater Treatment Plant | Liquids | 125,000 | 141 | 6/27/22 | 5/31/23 | ND | 241,919 | 0 |
| | | Jackson, MI | Jackson Wastewater Treatment Plant | Solids | 90,000 | 173 | 4/19/22 | 5/30/23 | ND | 163,392 | 1,611 |
| | | Jenison, MI | Grandville Clean Water Plant | Solids | 75,000 | 75 | 12/9/22 | 5/31/23 | ND | 146,044 | 1,314 |
| | | Mt. Pleasant, MI | Mt. Pleasant WRRF | Liquids | 21,690 | 20 | 4/9/23 | 5/31/23 | ND | 4,309 | 0 |
| | | Traverse City, MI | Traverse City Regional Waste Water Treatment Plant | Liquids | 30,623 | 56 | 1/23/23 | 5/31/23 | ND | 32,584 | 0 |
| | | Warren, MI | City of Warren Wastewater Treatment Plant | Liquids | 140,000 | 104 | 9/27/22 | 5/31/23 | ND | 917,610 | 0 |
| Minnesota | HHS5 | Mankato, MN | City of Mankato Water Resource Recovery Facility (WRRF) | Solids (Settling w/ Imhoff | 70,000 | 116 | 8/29/22 | 5/31/23 | ND | 111,906 | 0 |

| State | HHS Region | Plant | Site Name | Sample Type | Population Served | Sample Count | Sample Start | Sample End | Min IAV (cp/g) | Max IAV (cp/g) | Med IAV (cp/g) |
|---|---|---|---|---|---|---|---|---|---|---|---|
| | | | | cones onsite) | | | | | | | |
| | | Red Wing, MN | Red Wing Wastewater Treatment Facility | Solids (Settling w/ Imhoff cones onsite) | 16,000 | 9 | 5/8/23 | 5/25/23 | ND | 0 | 0 |
| | | Rochester, MN | City Of Rochester MN Water Reclamation Plant | Solids | 120,000 | 89 | 11/4/22 | 5/31/23 | ND | 179,473 | 1,647 |
| | | St. Cloud, MN | St. Cloud Nutrient, Energy and Water Recovery Facility | Liquids | 120,000 | 26 | 4/3/23 | 5/31/23 | ND | 4,187 | 0 |
| Nevada | HHS9 | Las Vegas, NV | Clark County Water Reclamation District (CCWRD) Flamingo Water Resource Center (FWRC) | Liquids | 990,000 | 29 | 3/29/23 | 5/31/23 | ND | 10,104 | 0 |

| State | HHS Region | Plant | Site Name | Sample Type | Population Served | Sample Count | Sample Start | Sample End | Min IAV (cp/g) | Max IAV (cp/g) | Med IAV (cp/g) |
|---|---|---|---|---|---|---|---|---|---|---|---|
| New Hampshire | HHS1 | Dover, NH | City of Dover Wastewater Treatment Facility | Liquids | 30,000 | 70 | 11/28/22 | 5/31/23 | ND | 290,177 | 3,969 |
| | | Hall Street, Concord, NH | Hall Street Wastewater Treatment Plant | Liquids | 45,000 | 99 | 10/12/22 | 5/31/23 | ND | 225,796 | 2,515 |
| | | Penacook, Concord, NH | Penacook Wastewater Treatment Facility | Liquids | 4,000 | 97 | 10/12/22 | 5/31/23 | ND | 585,759 | 0 |
| New Jersey | HHS2 | Belmar, NJ | South Monmouth Regional Sewerage Authority | Solids | 52,672 | 76 | 12/5/22 | 5/31/23 | ND | 114,789 | 0 |
| | | Bridgeton, NJ | Cumberland County Utilities Authority | Liquids | 50,000 | 32 | 3/13/23 | 5/31/23 | ND | 3,330 | 0 |
| | | Bridgewater, NJ | The Somerset Raritan Valley | Liquids | 130,000 | 8 | 5/15/23 | 5/31/23 | ND | 0 | 0 |

| State | HHS Region | Plant | Site Name | Sample Type | Population Served | Sample Count | Sample Start | Sample End | Min IAV (cp/g) | Max IAV (cp/g) | Med IAV (cp/g) |
|---|---|---|---|---|---|---|---|---|---|---|---|
| | | | Sewerage Authority | | | | | | | | |
| | | Newark, NJ | Passaic Valley Sewerage Commission | Solids | 1,500,000 | 113 | 8/5/22 | 5/31/23 | ND | 324,846 | 5,364 |
| | | Oakhurst, NJ | Township of Ocean Sewerage Authority | Liquids | 50,000 | 15 | 4/11/23 | 5/30/23 | ND | 2,973 | 0 |
| | | Union Beach, NJ | Bayshore Regional Sewerage Authority | Solids | 100,000 | 15 | 4/28/23 | 5/31/23 | ND | 4,185 | 0 |
| North Carolina | HHS4 | Kinston, NC | Johnnie Mosley Regional Water Reclamation Facility | Liquids | 25,000 | 83 | 10/27/22 | 5/4/23 | ND | 160,663 | 0 |
| | | Winston-Salem, NC | Archie Elledge WWTP | Liquids | 92,000 | 108 | 8/22/22 | 5/31/23 | ND | 123,596 | 0 |
| Ohio | HHS5 | Akron, OH | Akron Water Reclamation Facility | Solids | 365,000 | 55 | 1/6/23 | 5/31/23 | ND | 36,493 | 1,431 |

| State | HHS Region | Plant | Site Name | Sample Type | Population Served | Sample Count | Sample Start | Sample End | Min IAV (cp/g) | Max IAV (cp/g) | Med IAV (cp/g) |
|---|---|---|---|---|---|---|---|---|---|---|---|
| | | Youngstown, OH | City of Youngstown Wastewater Treatment Plant | Solids | 174,000 | 69 | 12/14/22 | 5/31/23 | ND | 131,445 | 1,906 |
| Pennsylvania | HHS3 | Chester, PA | DELCORA Western Regional Treatment Plant | Liquids | 220,000 | 80 | 10/30/22 | 5/30/23 | ND | 204,163 | 2,682 |
| | | Harrisburg, PA | Capital Region Water AWTF | Liquids | 125,000 | 130 | 8/2/22 | 5/30/23 | ND | 582,542 | 0 |
| South Carolina | HHS4 | Greenville, SC | Mauldin Road WWTP | Liquids | 142,376 | 2 | 8/17/22 | 8/19/22 | ND | 0 | 0 |
| South Dakota | HHS8 | Yankton, SD | City of Yankton Wastewater Treatment Facility | Liquids | 20,000 | 14 | 4/25/23 | 5/30/23 | ND | 4,191 | 0 |
| Texas | HHS6 | Gainesville, TX | City of Gainesville Wastewater Treatment Plant | Liquids | 17,300 | 58 | 12/22/22 | 5/31/23 | ND | 245,645 | 0 |

| State | HHS Region | Plant | Site Name | Sample Type | Population Served | Sample Count | Sample Start | Sample End | Min IAV (cp/g) | Max IAV (cp/g) | Med IAV (cp/g) |
|---|---|---|---|---|---|---|---|---|---|---|---|
| | | Garland, TX | City of Garland Rowlett Creek WWTP | Solids | 200,000 | 191 | 3/7/22 | 5/31/23 | ND | 190,007 | 0 |
| | | Hollywood Road, Amarillo, TX | Hollywood Road WWTP | Liquids | 60,000 | 73 | 12/7/22 | 5/30/23 | ND | 257,301 | 0 |
| | | River Road, Amarillo, TX | River Road WWTP | Liquids | 140,000 | 71 | 12/4/22 | 5/30/23 | ND | 464,466 | 0 |
| | | South, Laredo, TX | South Laredo WWTP | Liquids | 120,000 | 63 | 12/12/22 | 5/31/23 | ND | 44,178 | 0 |
| | | Sunnyvale, TX | Duck Creek Wastewater Treatment Plant | Solids | 186,000 | 171 | 2/27/22 | 5/4/23 | ND | 76,073 | 0 |
| | | Wichita Falls, TX | Wichita Falls Resource Recovery Facility | Solids | 90,000 | 75 | 12/5/22 | 5/31/23 | ND | 60,278 | 0 |
| | | Woodlands SJRA WWTF No. 1, TX | SJRA WWTF No.1 | Liquids | 65,000 | 44 | 2/20/23 | 5/31/23 | ND | 3,892 | 0 |

| State | HHS Region | Plant | Site Name | Sample Type | Population Served | Sample Count | Sample Start | Sample End | Min IAV (cp/g) | Max IAV (cp/g) | Med IAV (cp/g) |
|---|---|---|---|---|---|---|---|---|---|---|---|
| | | Woodlands SJRA WWTF No. 2, TX | SJRA WWTF No.2 | Liquids | 70,000 | 44 | 2/20/23 | 5/31/23 | ND | 5,837 | 0 |
| | | Woodlands SJRA WWTF No. 3, TX | SJRA WWTF No.3 | Liquids | 15,000 | 44 | 2/20/23 | 5/31/23 | ND | 14,318 | 0 |
| | | Zacate Creek, Laredo, TX | Zacate Creek WWTP | Liquids | 140,000 | 62 | 12/12/22 | 5/31/23 | ND | 149,949 | 0 |
| Utah | HHS8 | Central Salt Lake Valley, UT | Central Valley Water Reclamation Facility | Solids | 600,000 | 92 | 10/31/22 | 5/31/23 | ND | 272,992 | 4,770 |
| | | Provo, UT | Provo City Water Reclamation Facility | Solids (Settling w/ Imhoff cones onsite) | 115,000 | 73 | 9/19/22 | 5/31/23 | ND | 543,283 | 6,092 |
| Vermont | HHS1 | Essex Junction, VT | City of Essex Junction Wastewater Treatment Facility | Solids | 30,000 | 37 | 3/3/23 | 5/31/23 | ND | 12,630 | 0 |

| State | HHS Region | Plant | Site Name | Sample Type | Population Served | Sample Count | Sample Start | Sample End | Min IAV (cp/g) | Max IAV (cp/g) | Med IAV (cp/g) |
|---|---|---|---|---|---|---|---|---|---|---|---|
| | | Montpelier, VT | Montpelier Water Resource Recovery Facility | Solids | 10,100 | 37 | 3/6/23 | 5/30/23 | ND | 37,508 | 4,317 |
| | | South Burlington, VT | South Burlington-Airport Parkway WWTF | Liquids | 16,000 | 20 | 3/21/23 | 5/31/23 | ND | 7,854 | 0 |
| Virginia | HHS3 | Aquia, Stafford, VA | Aquia Wastewater Treatment Facility | Solids | 100,000 | 40 | 3/1/23 | 5/31/23 | ND | 4,418 | 0 |
| | | Hillsville, VA | Town of Hillsville Wastewater Treatment Plant | Solids | 3,000 | 72 | 12/12/22 | 5/30/23 | ND | 12,527 | 0 |
| | | Little Falls Run, Stafford, VA | Little Falls Run Wastewater Treatment Facility | Solids | 50,000 | 39 | 3/1/23 | 5/31/23 | ND | 3,122 | 0 |
| West Virginia | HHS3 | Wheeling, WV | City of Wheeling, Water Pollution Control Division | Solids | 100,000 | 23 | 4/10/23 | 3/1/23 | ND | 0 | 0 |

**Figure S3.** IAV concentration (log10 cp/g y-axis) by WWTP collected over the period of analysis of 6/1/2022 - 5/31/2023.

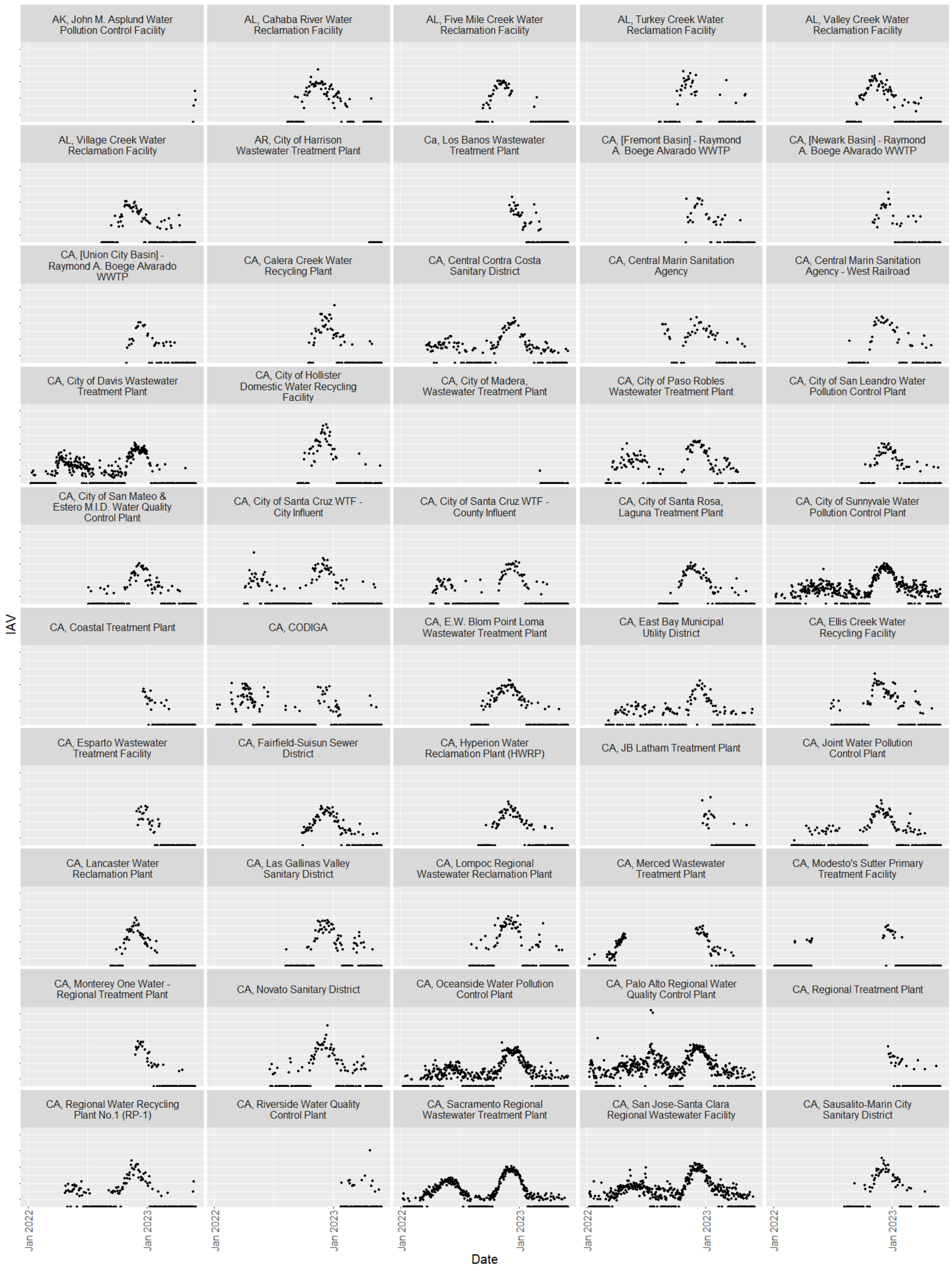
AK, John M. Asplund Water Pollution Control Facility
AL, Cahaba River Water Reclamation Facility
AL, Five Mile Creek Water Reclamation Facility
AL, Turkey Creek Water Reclamation Facility
AL, Valley Creek Water Reclamation Facility
AL, Village Creek Water Reclamation Facility
AR, City of Harrison Wastewater Treatment Plant
Ca, Los Banos Wastewater Treatment Plant
CA, [Fremont Basin] - Raymond A. Boege Alvarado WWTP
CA, [Newark Basin] - Raymond A. Boege Alvarado WWTP
CA, [Union City Basin] - Raymond A. Boege Alvarado WWTP
CA, Calera Creek Water Recycling Plant
CA, Central Contra Costa Sanitary District
CA, Central Marin Sanitation Agency
CA, Central Marin Sanitation Agency - West Railroad
CA, City of Davis Wastewater Treatment Plant
CA, City of Hollister Domestic Water Recycling Facility
CA, City of Madera, Wastewater Treatment Plant
CA, City of Paso Robles Wastewater Treatment Plant
CA, City of San Leandro Water Pollution Control Plant
CA, City of San Mateo & Estero M.I.D. Water Quality Control Plant
CA, City of Santa Cruz WTF - City Influent
CA, City of Santa Cruz WTF - County Influent
CA, City of Santa Rosa, Laguna Treatment Plant
CA, City of Sunnyvale Water Pollution Control Plant
CA, Coastal Treatment Plant
CA, CODIGA
CA, E.W. Blom Point Loma Wastewater Treatment Plant
CA, East Bay Municipal Utility District
CA, Ellis Creek Water Recycling Facility
CA, Esparto Wastewater Treatment Facility
CA, Fairfield-Suisun Sewer District
CA, Hyperion Water Reclamation Plant (HWRP)
CA, JB Latham Treatment Plant
CA, Joint Water Pollution Control Plant
CA, Lancaster Water Reclamation Plant
CA, Las Gallinas Valley Sanitary District
CA, Lompoc Regional Wastewater Reclamation Plant
CA, Merced Wastewater Treatment Plant
CA, Modesto's Sutter Primary Treatment Facility
CA, Monterey One Water - Regional Treatment Plant
CA, Novato Sanitary District
CA, Oceanside Water Pollution Control Plant
CA, Palo Alto Regional Water Quality Control Plant
CA, Regional Treatment Plant
CA, Regional Water Recycling Plant No.1 (RP-1)
CA, Riverside Water Quality Control Plant
CA, Sacramento Regional Wastewater Treatment Plant
CA, San Jose-Santa Clara Regional Wastewater Facility
CA, Sausalito-Marin City Sanitary District
IAV
Jan 2022
Jan 2023
Date

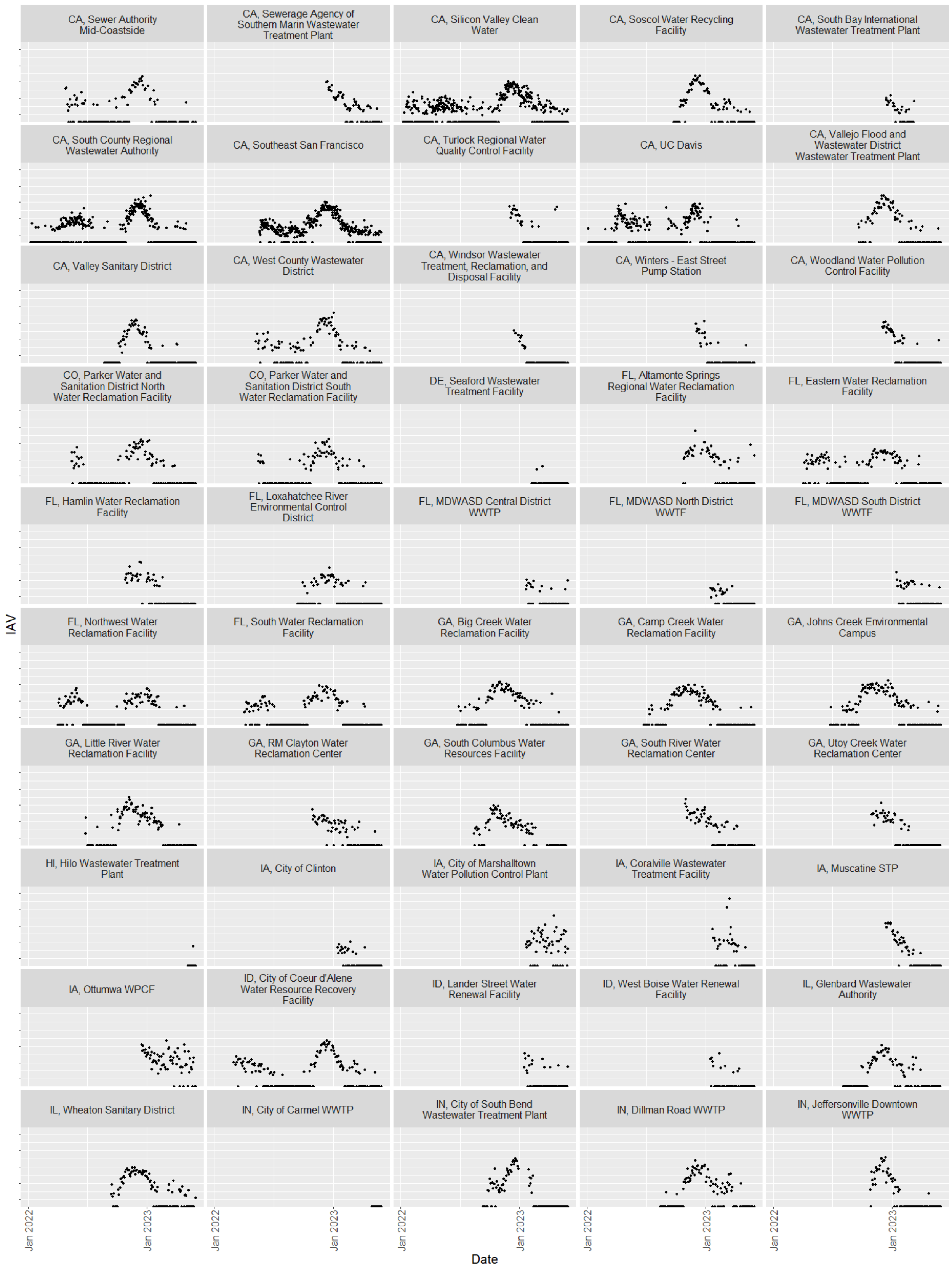
CA, Sewer Authority Mid-Coastside
CA, Sewerage Agency of Southern Marin Wastewater Treatment Plant
CA, Silicon Valley Clean Water
CA, Soscol Water Recycling Facility
CA, South Bay International Wastewater Treatment Plant
CA, South County Regional Wastewater Authority
CA, Southeast San Francisco
CA, Turlock Regional Water Quality Control Facility
CA, UC Davis
CA, Vallejo Flood and Wastewater District Wastewater Treatment Plant
CA, Valley Sanitary District
CA, West County Wastewater District
CA, Windsor Wastewater Treatment, Reclamation, and Disposal Facility
CA, Winters - East Street Pump Station
CA, Woodland Water Pollution Control Facility
CO, Parker Water and Sanitation District North Water Reclamation Facility
CO, Parker Water and Sanitation District South Water Reclamation Facility
DE, Seaford Wastewater Treatment Facility
FL, Altamonte Springs Regional Water Reclamation Facility
FL, Eastern Water Reclamation Facility
FL, Hamlin Water Reclamation Facility
FL, Loxahatchee River Environmental Control District
FL, MDWASD Central District WWTP
FL, MDWASD North District WWTF
FL, MDWASD South District WWTF
FL, Northwest Water Reclamation Facility
FL, South Water Reclamation Facility
GA, Big Creek Water Reclamation Facility
GA, Camp Creek Water Reclamation Facility
GA, Johns Creek Environmental Campus
GA, Little River Water Reclamation Facility
GA, RM Clayton Water Reclamation Center
GA, South Columbus Water Resources Facility
GA, South River Water Reclamation Center
GA, Utoy Creek Water Reclamation Center
HI, Hilo Wastewater Treatment Plant
IA, City of Clinton
IA, City of Marshalltown Water Pollution Control Plant
IA, Coralville Wastewater Treatment Facility
IA, Muscatine STP
IA, Ottumwa WPCF
ID, City of Coeur d'Alene Water Resource Recovery Facility
ID, Lander Street Water Renewal Facility
ID, West Boise Water Renewal Facility
IL, Glenbard Wastewater Authority
IL, Wheaton Sanitary District
IN, City of Carmel WWTP
IN, City of South Bend Wastewater Treatment Plant
IN, Dillman Road WWTP
IN, Jeffersonville Downtown WWTP
IAV
Jan 2022
Jan 2023
Date

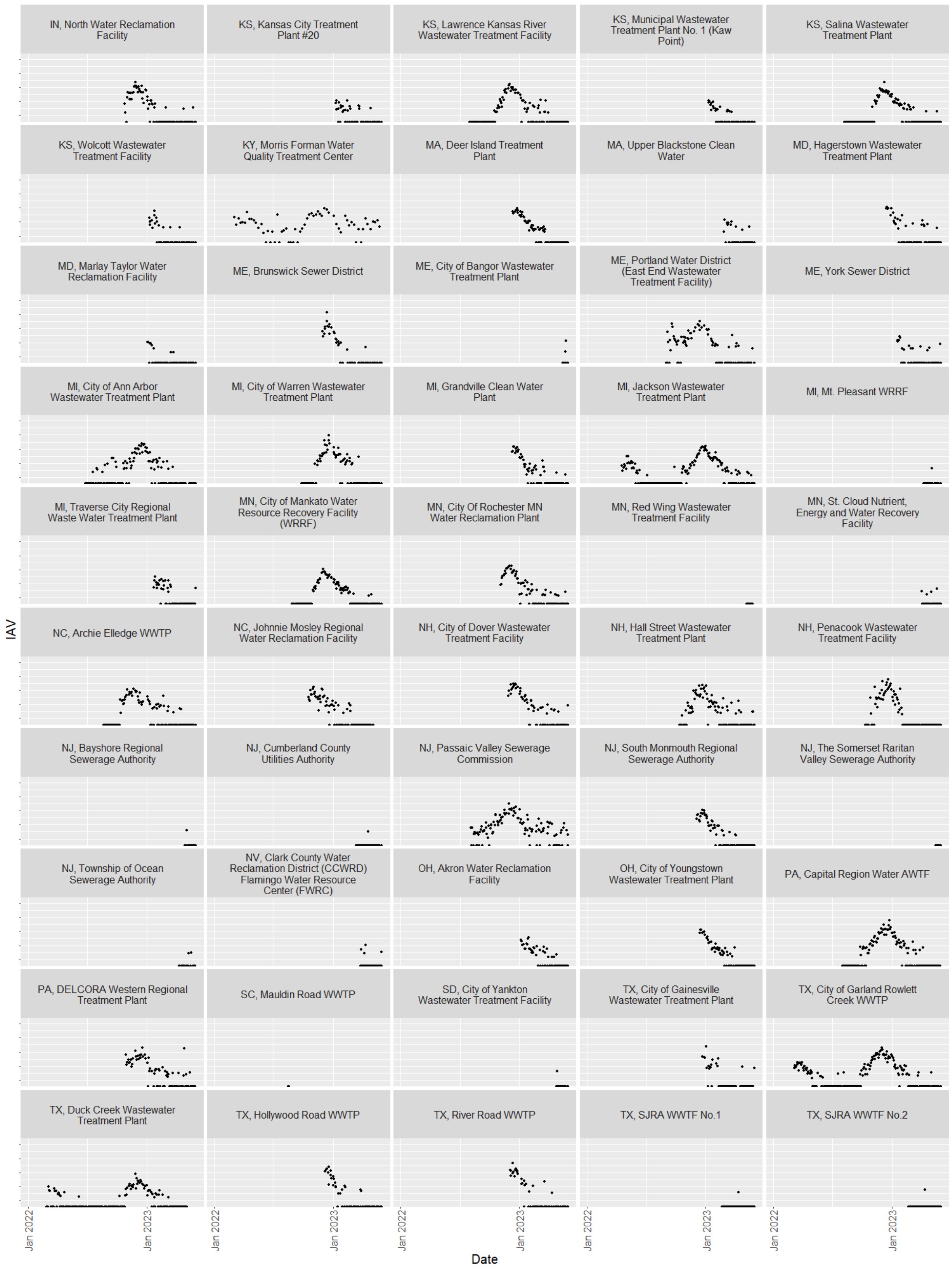
IN, North Water Reclamation Facility
KS, Kansas City Treatment Plant #20
KS, Lawrence Kansas River Wastewater Treatment Facility
KS, Municipal Wastewater Treatment Plant No. 1 (Kaw Point)
KS, Salina Wastewater Treatment Plant
KS, Wolcott Wastewater Treatment Facility
KY, Morris Forman Water Quality Treatment Center
MA, Deer Island Treatment Plant
MA, Upper Blackstone Clean Water
MD, Hagerstown Wastewater Treatment Plant
MD, Marlay Taylor Water Reclamation Facility
ME, Brunswick Sewer District
ME, City of Bangor Wastewater Treatment Plant
ME, Portland Water District (East End Wastewater Treatment Facility)
ME, York Sewer District
MI, City of Ann Arbor Wastewater Treatment Plant
MI, City of Warren Wastewater Treatment Plant
MI, Grandville Clean Water Plant
MI, Jackson Wastewater Treatment Plant
MI, Mt. Pleasant WRRF
MI, Traverse City Regional Waste Water Treatment Plant
MN, City of Mankato Water Resource Recovery Facility (WRRF)
MN, City Of Rochester MN Water Reclamation Plant
MN, Red Wing Wastewater Treatment Facility
MN, St. Cloud Nutrient, Energy and Water Recovery Facility
NC, Archie Elledge WWTP
NC, Johnnie Mosley Regional Water Reclamation Facility
NH, City of Dover Wastewater Treatment Facility
NH, Hall Street Wastewater Treatment Plant
NH, Penacook Wastewater Treatment Facility
NJ, Bayshore Regional Sewerage Authority
NJ, Cumberland County Utilities Authority
NJ, Passaic Valley Sewerage Commission
NJ, South Monmouth Regional Sewerage Authority
NJ, The Somerset Raritan Valley Sewerage Authority
NJ, Township of Ocean Sewerage Authority
NV, Clark County Water Reclamation District (CCWRD) Flamingo Water Resource Center (FWRC)
OH, Akron Water Reclamation Facility
OH, City of Youngstown Wastewater Treatment Plant
PA, Capital Region Water AWTF
PA, DELCORA Western Regional Treatment Plant
SC, Mauldin Road WWTP
SD, City of Yankton Wastewater Treatment Facility
TX, City of Gainesville Wastewater Treatment Plant
TX, City of Garland Rowlett Creek WWTP
TX, Duck Creek Wastewater Treatment Plant
TX, Hollywood Road WWTP
TX, River Road WWTP
TX, SJRA WWTF No.1
TX, SJRA WWTF No.2
IAV
Jan 2022
Jan 2023
Date

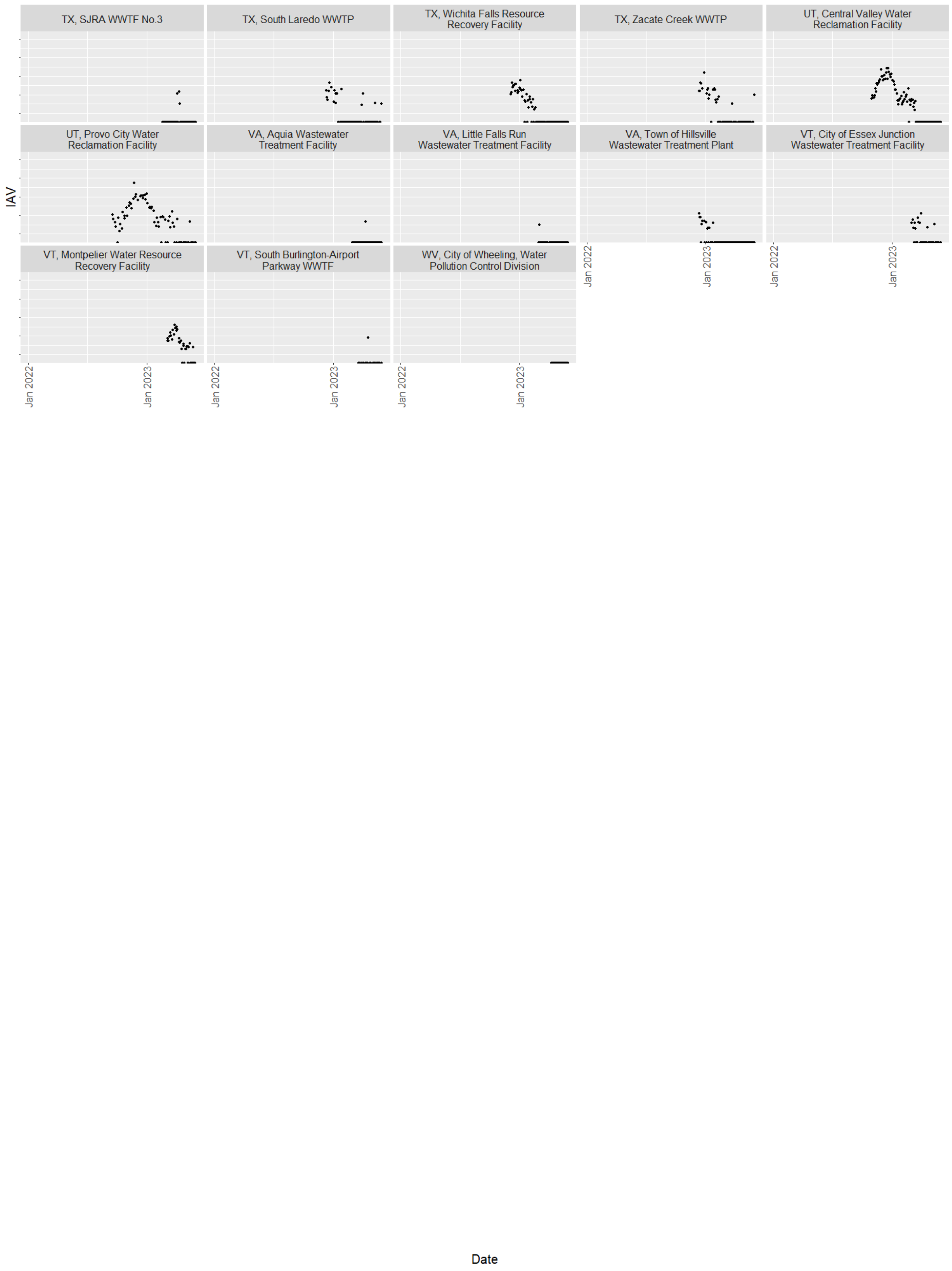
TX, SJRA WWTF No.3
TX, South Laredo WWTP
TX, Wichita Falls Resource Recovery Facility
TX, Zacate Creek WWTP
UT, Central Valley Water Reclamation Facility
UT, Provo City Water Reclamation Facility
VA, Aquia Wastewater Treatment Facility
VA, Little Falls Run Wastewater Treatment Facility
VA, Town of Hillsville Wastewater Treatment Plant
VT, City of Essex Junction Wastewater Treatment Facility
VT, Montpelier Water Resource Recovery Facility
VT, South Burlington-Airport Parkway WWTF
WV, City of Wheeling, Water Pollution Control Division
IAV
Jan 2022
Jan 2023
Date

**Figure S4.** IAV/PMMoV (log10 IAV/PMMoV y-axis) by WWTP collected over the period of analysis of 6/1/2022 - 5/31/2023

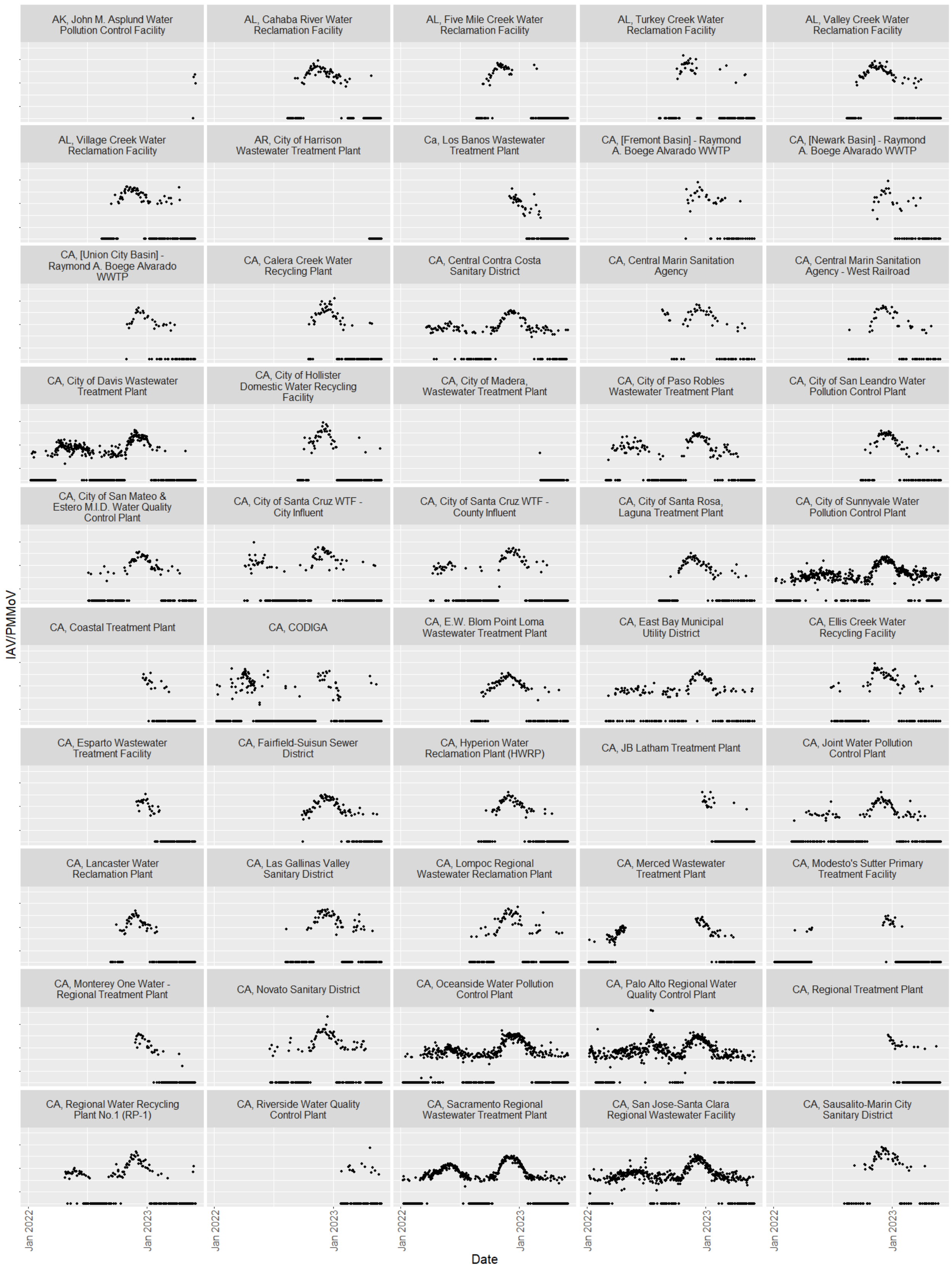
AK, John M. Asplund Water Pollution Control Facility
AL, Cahaba River Water Reclamation Facility
AL, Five Mile Creek Water Reclamation Facility
AL, Turkey Creek Water Reclamation Facility
AL, Valley Creek Water Reclamation Facility
AL, Village Creek Water Reclamation Facility
AR, City of Harrison Wastewater Treatment Plant
Ca, Los Banos Wastewater Treatment Plant
CA, [Fremont Basin] - Raymond A. Boege Alvarado WWTP
CA, [Newark Basin] - Raymond A. Boege Alvarado WWTP
CA, [Union City Basin] - Raymond A. Boege Alvarado WWTP
CA, Calera Creek Water Recycling Plant
CA, Central Contra Costa Sanitary District
CA, Central Marin Sanitation Agency
CA, Central Marin Sanitation Agency - West Railroad
CA, City of Davis Wastewater Treatment Plant
CA, City of Hollister Domestic Water Recycling Facility
CA, City of Madera, Wastewater Treatment Plant
CA, City of Paso Robles Wastewater Treatment Plant
CA, City of San Leandro Water Pollution Control Plant
CA, City of San Mateo & Estero M.I.D. Water Quality Control Plant
CA, City of Santa Cruz WTF - City Influent
CA, City of Santa Cruz WTF - County Influent
CA, City of Santa Rosa, Laguna Treatment Plant
CA, City of Sunnyvale Water Pollution Control Plant
CA, Coastal Treatment Plant
CA, CODIGA
CA, E.W. Blom Point Loma Wastewater Treatment Plant
CA, East Bay Municipal Utility District
CA, Ellis Creek Water Recycling Facility
CA, Esparto Wastewater Treatment Facility
CA, Fairfield-Suisun Sewer District
CA, Hyperion Water Reclamation Plant (HWRP)
CA, JB Latham Treatment Plant
CA, Joint Water Pollution Control Plant
CA, Lancaster Water Reclamation Plant
CA, Las Gallinas Valley Sanitary District
CA, Lompoc Regional Wastewater Reclamation Plant
CA, Merced Wastewater Treatment Plant
CA, Modesto's Sutter Primary Treatment Facility
CA, Monterey One Water - Regional Treatment Plant
CA, Novato Sanitary District
CA, Oceanside Water Pollution Control Plant
CA, Palo Alto Regional Water Quality Control Plant
CA, Regional Treatment Plant
CA, Regional Water Recycling Plant No.1 (RP-1)
CA, Riverside Water Quality Control Plant
CA, Sacramento Regional Wastewater Treatment Plant
CA, San Jose-Santa Clara Regional Wastewater Facility
CA, Sausalito-Marin City Sanitary District
IAV/PMMoV
Jan 2022
Jan 2023
Date

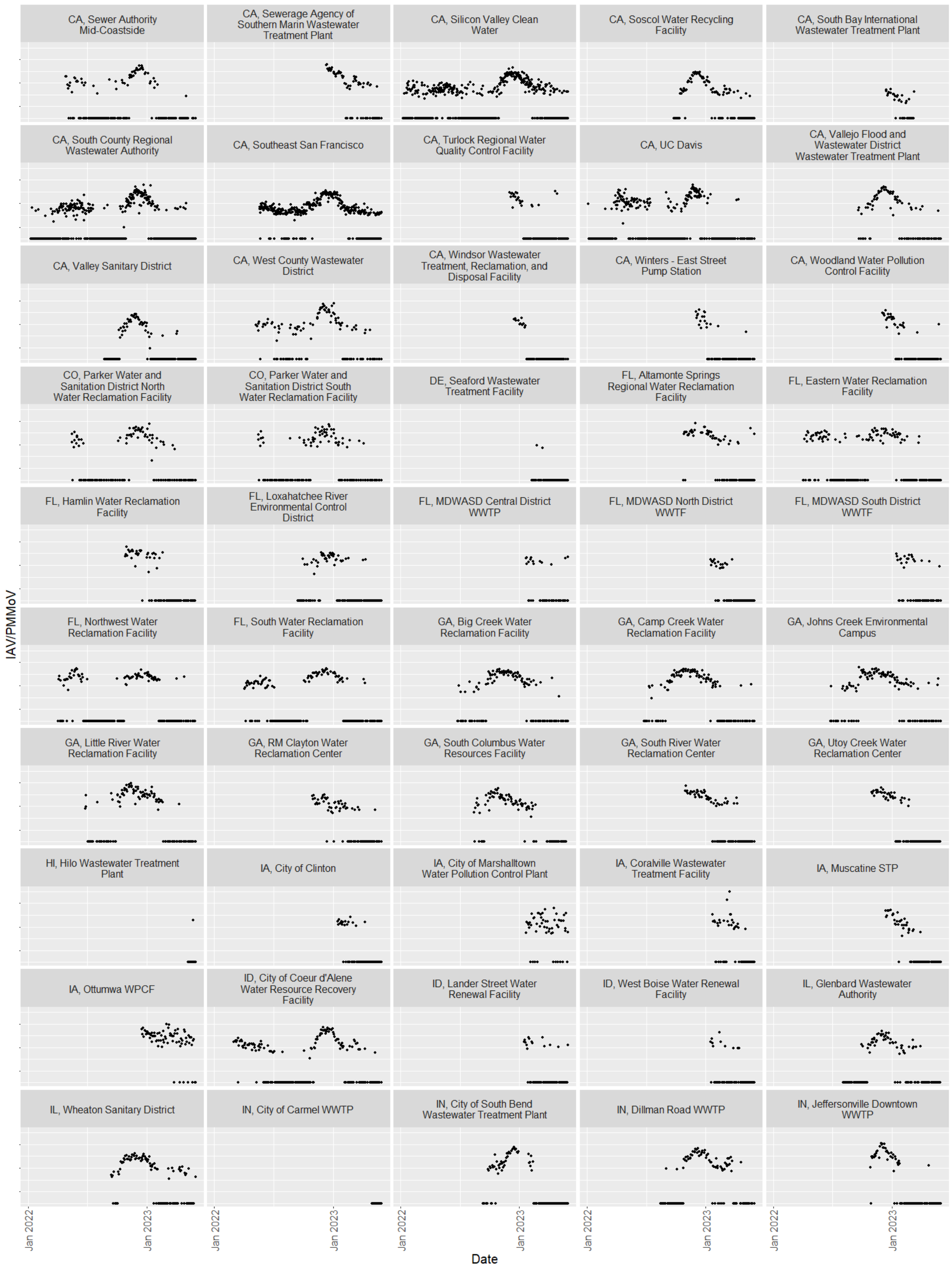
CA, Sewer Authority Mid-Coastside
CA, Sewerage Agency of Southern Marin Wastewater Treatment Plant
CA, Silicon Valley Clean Water
CA, Soscol Water Recycling Facility
CA, South Bay International Wastewater Treatment Plant
CA, South County Regional Wastewater Authority
CA, Southeast San Francisco
CA, Turlock Regional Water Quality Control Facility
CA, UC Davis
CA, Vallejo Flood and Wastewater District Wastewater Treatment Plant
CA, Valley Sanitary District
CA, West County Wastewater District
CA, Windsor Wastewater Treatment, Reclamation, and Disposal Facility
CA, Winters - East Street Pump Station
CA, Woodland Water Pollution Control Facility
CO, Parker Water and Sanitation District North Water Reclamation Facility
CO, Parker Water and Sanitation District South Water Reclamation Facility
DE, Seaford Wastewater Treatment Facility
FL, Altamonte Springs Regional Water Reclamation Facility
FL, Eastern Water Reclamation Facility
FL, Hamlin Water Reclamation Facility
FL, Loxahatchee River Environmental Control District
FL, MDWASD Central District WWTP
FL, MDWASD North District WWTF
FL, MDWASD South District WWTF
FL, Northwest Water Reclamation Facility
FL, South Water Reclamation Facility
GA, Big Creek Water Reclamation Facility
GA, Camp Creek Water Reclamation Facility
GA, Johns Creek Environmental Campus
GA, Little River Water Reclamation Facility
GA, RM Clayton Water Reclamation Center
GA, South Columbus Water Resources Facility
GA, South River Water Reclamation Center
GA, Utoy Creek Water Reclamation Center
HI, Hilo Wastewater Treatment Plant
IA, City of Clinton
IA, City of Marshalltown Water Pollution Control Plant
IA, Coralville Wastewater Treatment Facility
IA, Muscatine STP
IA, Ottumwa WPCF
ID, City of Coeur d'Alene Water Resource Recovery Facility
ID, Lander Street Water Renewal Facility
ID, West Boise Water Renewal Facility
IL, Glenbard Wastewater Authority
IL, Wheaton Sanitary District
IN, City of Carmel WWTP
IN, City of South Bend Wastewater Treatment Plant
IN, Dillman Road WWTP
IN, Jeffersonville Downtown WWTP
IAV/PMMoV
Jan 2022
Jan 2023
Date

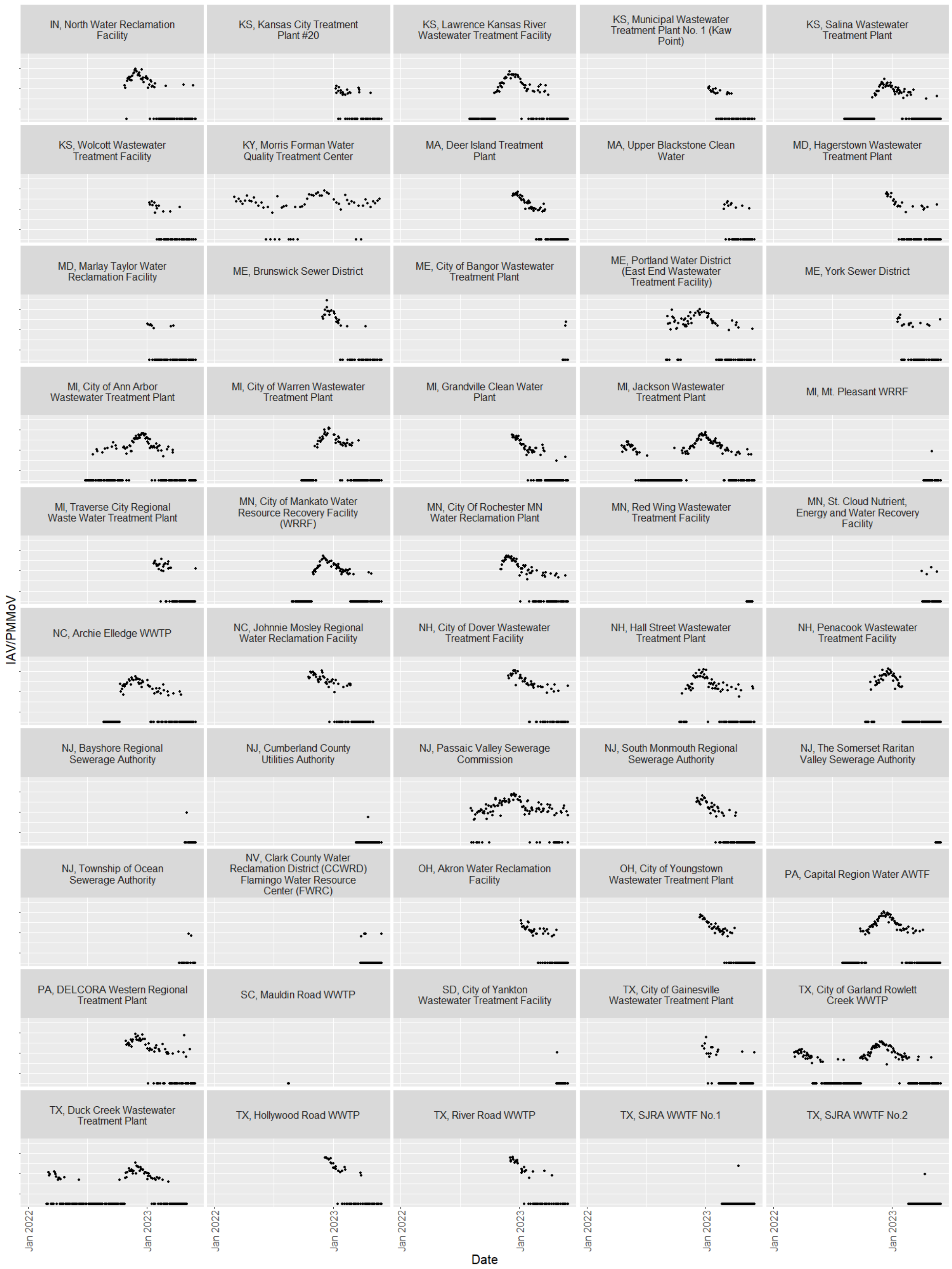
IN, North Water Reclamation Facility
KS, Kansas City Treatment Plant #20
KS, Lawrence Kansas River Wastewater Treatment Facility
KS, Municipal Wastewater Treatment Plant No. 1 (Kaw Point)
KS, Salina Wastewater Treatment Plant
KS, Wolcott Wastewater Treatment Facility
KY, Morris Forman Water Quality Treatment Center
MA, Deer Island Treatment Plant
MA, Upper Blackstone Clean Water
MD, Hagerstown Wastewater Treatment Plant
MD, Marlay Taylor Water Reclamation Facility
ME, Brunswick Sewer District
ME, City of Bangor Wastewater Treatment Plant
ME, Portland Water District (East End Wastewater Treatment Facility)
ME, York Sewer District
MI, City of Ann Arbor Wastewater Treatment Plant
MI, City of Warren Wastewater Treatment Plant
MI, Grandville Clean Water Plant
MI, Jackson Wastewater Treatment Plant
MI, Mt. Pleasant WRRF
MI, Traverse City Regional Waste Water Treatment Plant
MN, City of Mankato Water Resource Recovery Facility (WRRF)
MN, City Of Rochester MN Water Reclamation Plant
MN, Red Wing Wastewater Treatment Facility
MN, St. Cloud Nutrient, Energy and Water Recovery Facility
NC, Archie Elledge WWTP
NC, Johnnie Mosley Regional Water Reclamation Facility
NH, City of Dover Wastewater Treatment Facility
NH, Hall Street Wastewater Treatment Plant
NH, Penacook Wastewater Treatment Facility
NJ, Bayshore Regional Sewerage Authority
NJ, Cumberland County Utilities Authority
NJ, Passaic Valley Sewerage Commission
NJ, South Monmouth Regional Sewerage Authority
NJ, The Somerset Raritan Valley Sewerage Authority
NJ, Township of Ocean Sewerage Authority
NV, Clark County Water Reclamation District (CCWRD) Flamingo Water Resource Center (FWRC)
OH, Akron Water Reclamation Facility
OH, City of Youngstown Wastewater Treatment Plant
PA, Capital Region Water AWTF
PA, DELCORA Western Regional Treatment Plant
SC, Mauldin Road WWTP
SD, City of Yankton Wastewater Treatment Facility
TX, City of Gainesville Wastewater Treatment Plant
TX, City of Garland Rowlett Creek WWTP
TX, Duck Creek Wastewater Treatment Plant
TX, Hollywood Road WWTP
TX, River Road WWTP
TX, SJRA WWTF No.1
TX, SJRA WWTF No.2
IAV/PMMoV
Jan 2022
Jan 2023
Date

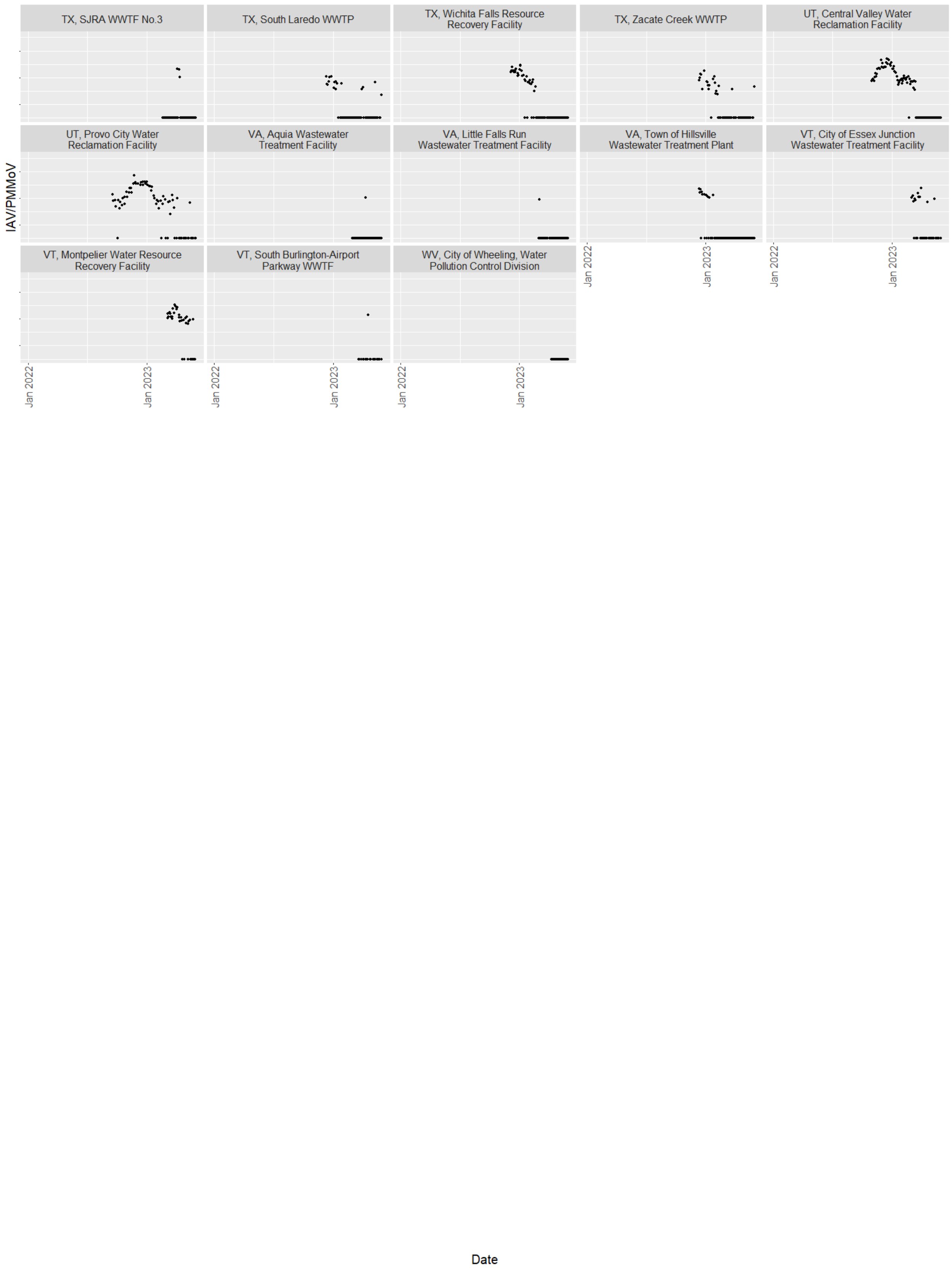

TX, SJRA WWTF No.3
TX, South Laredo WWTP
TX, Wichita Falls Resource Recovery Facility
TX, Zacate Creek WWTP
UT, Central Valley Water Reclamation Facility
UT, Provo City Water Reclamation Facility
VA, Aquia Wastewater Treatment Facility
VA, Little Falls Run Wastewater Treatment Facility
VA, Town of Hillsville Wastewater Treatment Plant
VT, City of Essex Junction Wastewater Treatment Facility
VT, Montpelier Water Resource Recovery Facility
VT, South Burlington-Airport Parkway WWTF
WV, City of Wheeling, Water Pollution Control Division
IAV/PMMoV
Jan 2022
Jan 2023
Date

**Table S2.** Avg_IAV_Region baseline values for HHS Regions. NA: not applicable due to less than one year of data at time of analysis. ?: the ILI offset had not yet occurred at the time of analysis

| HHS Region | Description | ILI onset | ILI offset | IAV/PMMoV mean | IAV/PMMoV Geometric mean | IAV/PMMoV 2 x min |
|---|---|---|---|---|---|---|
| 1 | Connecticut, Maine, Massachusetts, New Hampshire, Rhode Island, and Vermont | $2.73 \times 10^{-5}$ | $6.90 \times 10^{-5}$ | NA | NA | $3.46 \times 10^{-6}$ |
| 4 | Alabama, Florida, Georgia, Kentucky, Mississippi, North Carolina, South Carolina, and Tennessee | $2.04 \times 10^{-5}$ | $3.60 \times 10^{-5}$ | $4.14 \times 10^{-5}$ | $1.74 \times 10^{-5}$ | $2.85 \times 10^{-6}$ |
| 5 | Illinois, Indiana, Michigan, Minnesota, Ohio, and Wisconsin | $5.58 \times 10^{-6}$ | $8.55 \times 10^{-5}$ | $4.06 \times 10^{-5}$ | $6.59 \times 10^{-6}$ | $2.11 \times 10^{-6}$ |
| 8 | Colorado, Montana, North Dakota, South Dakota, Utah, and Wyoming | $5.72 \times 10^{-6}$ | $8.55 \times 10^{-6}$ | $1.88 \times 10^{-5}$ | $3.78 \times 10^{-6}$ | $9.19 \times 10^{-7}$ |

| HHS Region | Description | ILI onset | ILI offset | IAV/PMMoV mean | IAV/PMMoV Geometric mean | IAV/PMMoV 2 x min |
|---|---|---|---|---|---|---|
| 9 | Arizona, California, Hawaii, and Nevada | $2.31 \times 10^{-6}$ | ? | $1.06 \times 10^{-5}$ | $3.70 \times 10^{-6}$ | $5.70 \times 10^{-7}$ |

**Table S3.** Avg_IAV_State baseline values for states. NA: not applicable due to less than one year of data at time of analysis.

| State | IAV/PMMoV mean | IAV/PMMoV Geometric mean | IAV/PMMoV 2 x min |
|---|---|---|---|
| CA | $1.02 \times 10^{-5}$ | $2.77 \times 10^{-6}$ | $8.10 \times 10^{-7}$ |
| FL | $3.00 \times 10^{-5}$ | $1.01 \times 10^{-5}$ | $3.54 \times 10^{-6}$ |
| GA | $3.11 \times 10^{-5}$ | $7.60 \times 10^{-6}$ | $1.78 \times 10^{-6}$ |
| MI | $5.12 \times 10^{-5}$ | $6.38 \times 10^{-6}$ | $2.26 \times 10^{-6}$ |
| TX | $1.20 \times 10^{-5}$ | $1.90 \times 10^{-6}$ | $9.44 \times 10^{-7}$ |
| AL | NA | NA | $2.50 \times 10^{-6}$ |
| IL | NA | NA | $1.55 \times 10^{-6}$ |
| KS | NA | NA | $1.40 \times 10^{-6}$ |
| NH | NA | NA | $3.25 \times 10^{-6}$ |
| CO | NA | NA | $3.54 \times 10^{-6}$ |

**Table S4.** Influenza season onset, offset, and duration for 2022-2023 influenza season for selected HHS regions based on the ILI baseline (ordered by ILI onset) or calculated using Avg_IAV_Region mean, geometric mean, or twice the minimum observation.

ND: Not determined due to less than 1 year of data. NA: baseline value exceeded in all previous observations. *: the offset had not yet occurred at the time of analysis.

| **Baseline** | **HHS Regions Onset/Offset (duration in days)** | | | | |
|---|---|---|---|---|---|
| | 4 | 9 | 5 | 8 | 1 |
| *ILI* | 9/26/2022-1/15/2023 (111) | 10/9/2022-* (*) | 10/16/2022-1/8/2023 (84) | 10/23/2022-1/22/2023 (91) | 10/23/2022-1/15/2023 (84) |
| *IAV/PMMoV mean* | 10/6/2022-1/10/2023 (96) | 10/31/2022-1/14/2023 (75) | 11/14/2022-1/22/2023 (69) | 11/9/2022-1/17/2023 (69) | ND |

| | | | | | |
|---|---|---|---|---|---|
| *IAV/PMMoV geometric mean* | 9/24/2022-1/29/2023 (127) | 10/15/2022-1/24/2023 (101) | 10/19/2022-2/23/2023 (127) | 10/19/2022-3/8/2023 (140) | ND |
| *IAV/PMMoV 2x min* | 7/13/2022-* (*) | NA-4/28/2023 (NA) | 9/16/2022-3/29/2023 (194) | 9/19/2022-3/22/2023 (184) | NA-3/24/2023 (NA) |

**Table S5**. Flu season onset, offset, and duration for states calculated using Avg_IAV_State mean, geometric mean, or twice the minimum observation. ND: not determined due to less than 1 year of data. NA: baseline value exceeded in all previous observations.

| State | Onset | | | Offset | | | Duration (days) | | |
|---|---|---|---|---|---|---|---|---|---|
| | **2 x min** | **geomean** | **mean** | **2 x min** | **geomean** | **mean** | **2 x min** | **geomean** | **mean** |
| CA | NA | 10/11/22 | 11/1/22 | 3/28/23 | 1/26/23 | 1/14/23 | NA | 107 | 74 |
| FL | 9/25/22 | 10/16/22 | 10/21/22 | 3/14/23 | 3/8/23 | 1/22/23 | 170 | 143 | 93 |
| GA | 8/13/22 | 9/17/22 | 9/21/22 | 3/11/23 | 2/8/23 | 1/8/23 | 210 | 144 | 109 |
| MI | 10/18/22 | 11/6/22 | 11/19/22 | 3/20/23 | 3/4/23 | 1/19/23 | 153 | 118 | 61 |

| | | | | | | | | | |
|---|---|---|---|---|---|---|---|---|---|
| TX | 9/27/22 | 9/30/22 | 10/29/22 | 2/18/23 | 2/12/23 | 1/9/23 | 144 | 135 | 72 |
| AL | 9/18/22 | ND | ND | 2/23/23 | ND | ND | 158 | ND | ND |
| CO | 10/25/22 | ND | ND | 2/28/23 | ND | ND | 126 | ND | ND |
| IL | 9/28/22 | ND | ND | 4/30/23 | ND | ND | 214 | ND | ND |
| KS | 10/19/22 | ND | ND | 3/7/23 | ND | ND | 139 | ND | ND |
| NH | 11/1/22 | ND | ND | 4/19/23 | ND | ND | 169 | ND | ND |

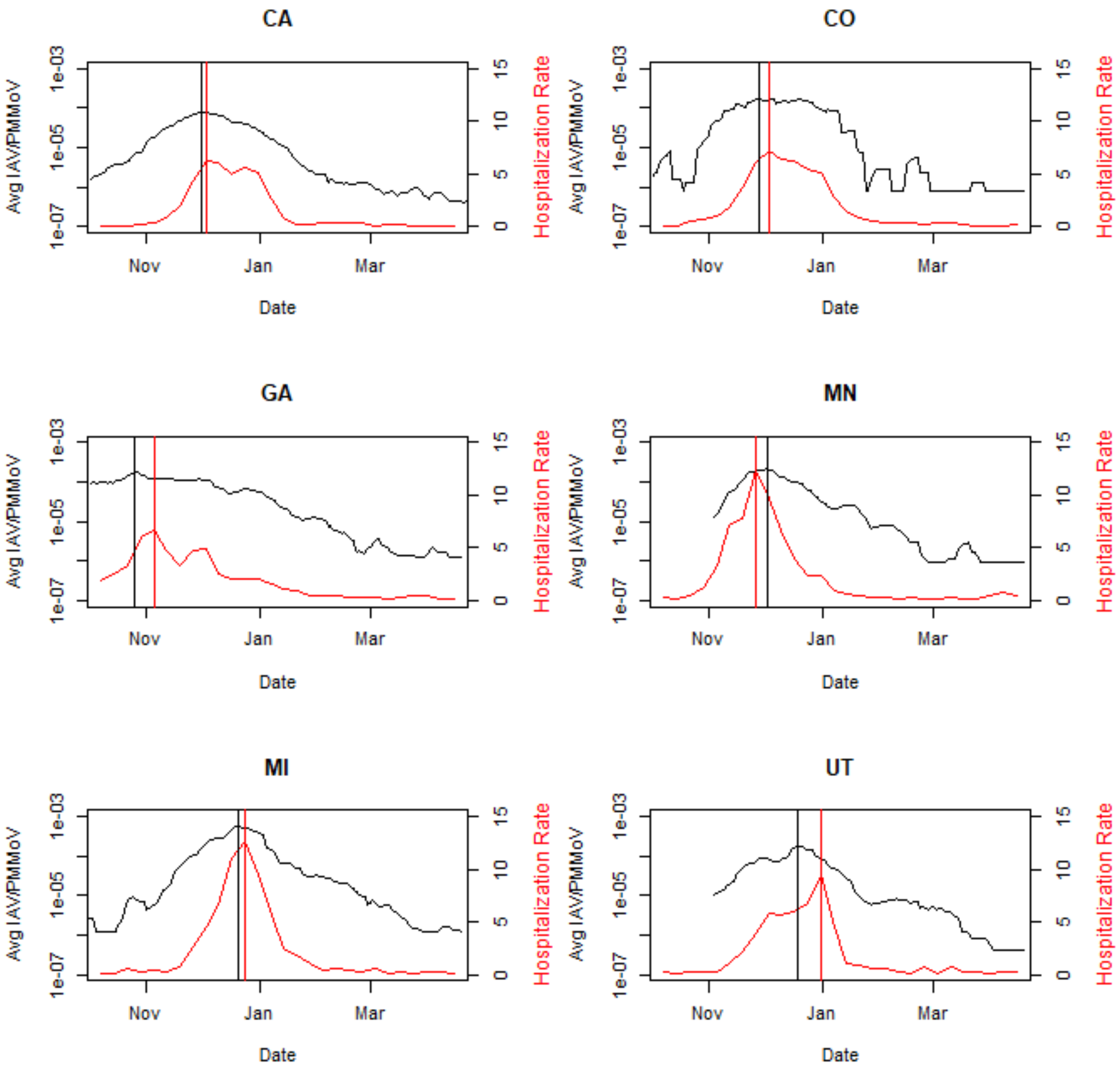

**Figure S5.** 2022-2023 influenza season peak (vertical line) and intensity (y-axis) by state using Avg_IAV_State or FluSurv-NET hospitalization rate (hospitalizations per 100,000 population) for states with both sets of data

**Table S6.** Comparison of peak and intensity using hospitalization rate and Avg_IAV_State by state ordered by hospitalization rate intensity.

| State | Hospitalization Rate | | IAV/PMMoV | |
|---|---|---|---|---|
| | Intensity | Peak | Intensity | Peak |
| CA | 6.3 | 12/3/22 | $7.9x10^{-5}$ | 12/1/22 |
| GA | 6.7 | 11/5/22 | $1.9x10^{-4}$ | 10/26/22 |
| CO | 7.2 | 12/3/22 | $1.8x10^{-4}$ | 11/28/22 |
| UT | 9.4 | 12/31/22 | $1.8x10^{-4}$ | 12/19/22 |
| MN | 12.4 | 11/26/22 | $2.1x10^{-4}$ | 12/2/22 |
| MI | 12.7 | 12/24/22 | $5.5x10^{-4}$ | 12/20/22 |

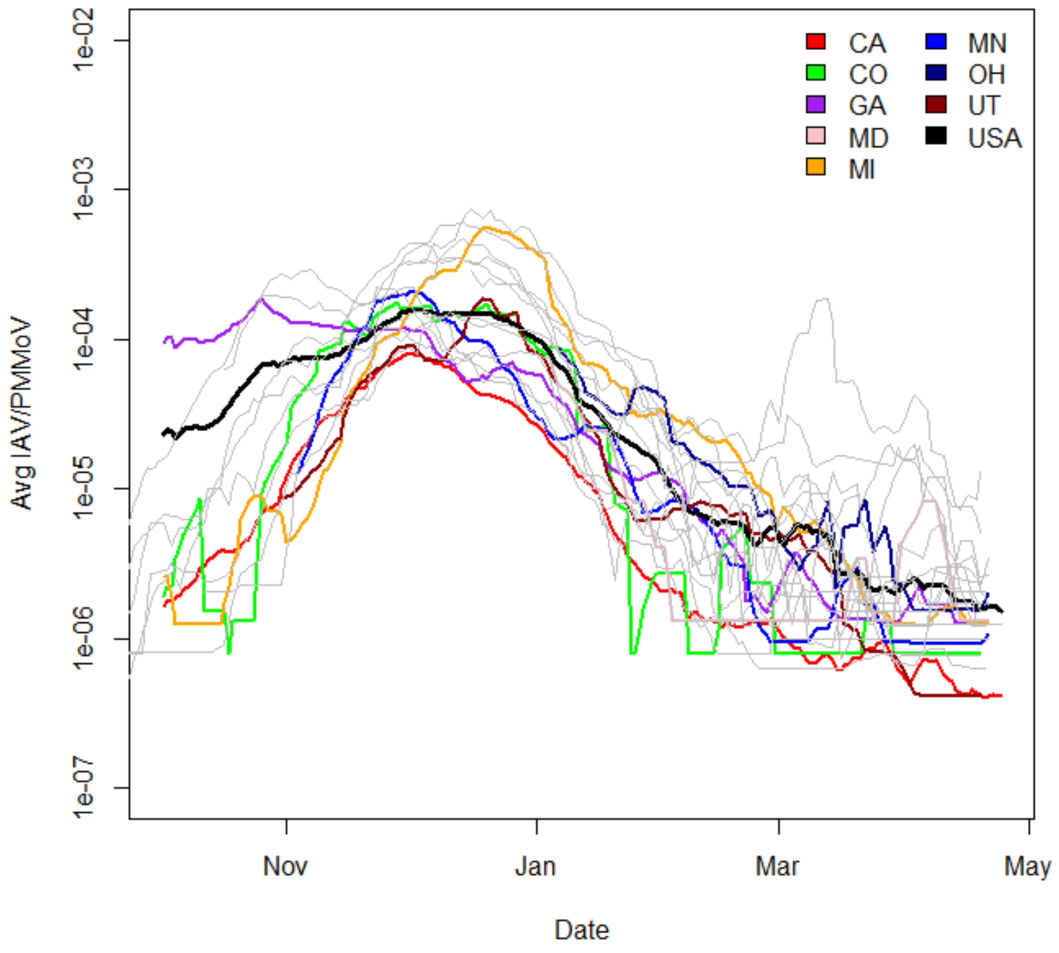

**Figure S6**. 2022-2023 flu Season Avg_IAV_State for all states. States in color share both wastewater and FluServe-Net data. States in grayscale only have wastewater.

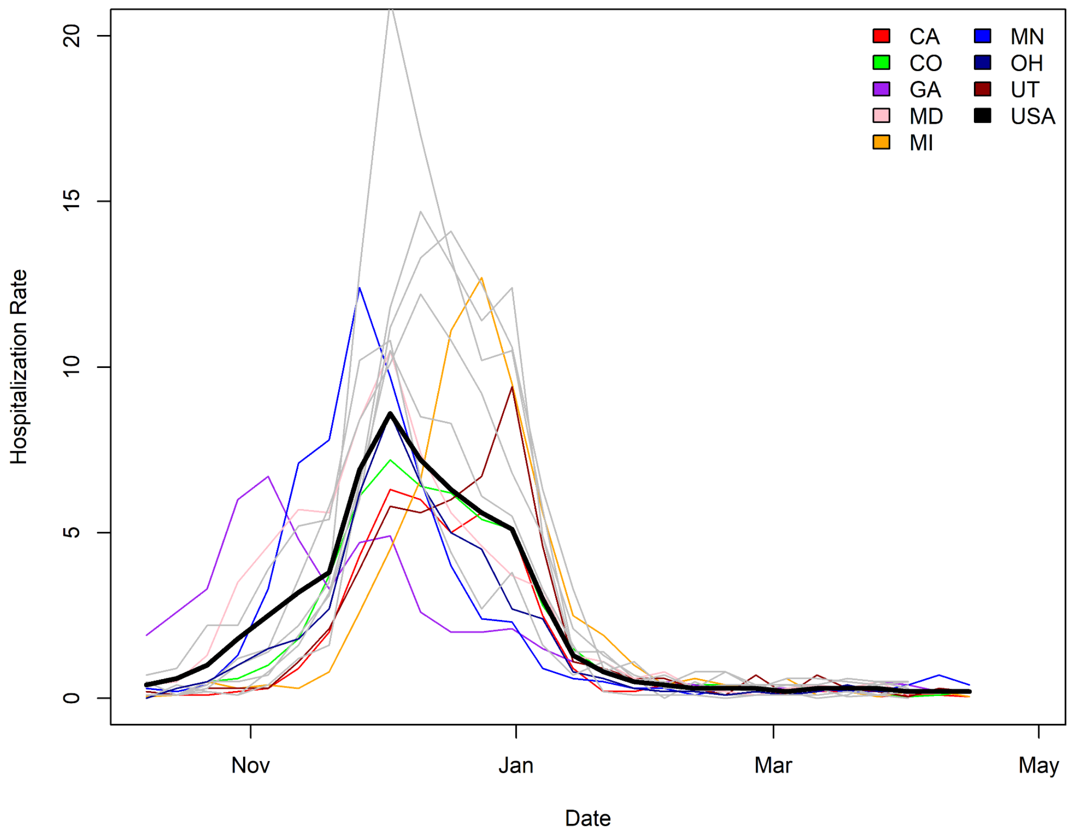

**Figure S7.** 2022-2023 flu season hospitalization rate for all FluServe-Net states. States in color share both wastewater and FluServe-Net data. States in grayscale only have FluServe-Net data.

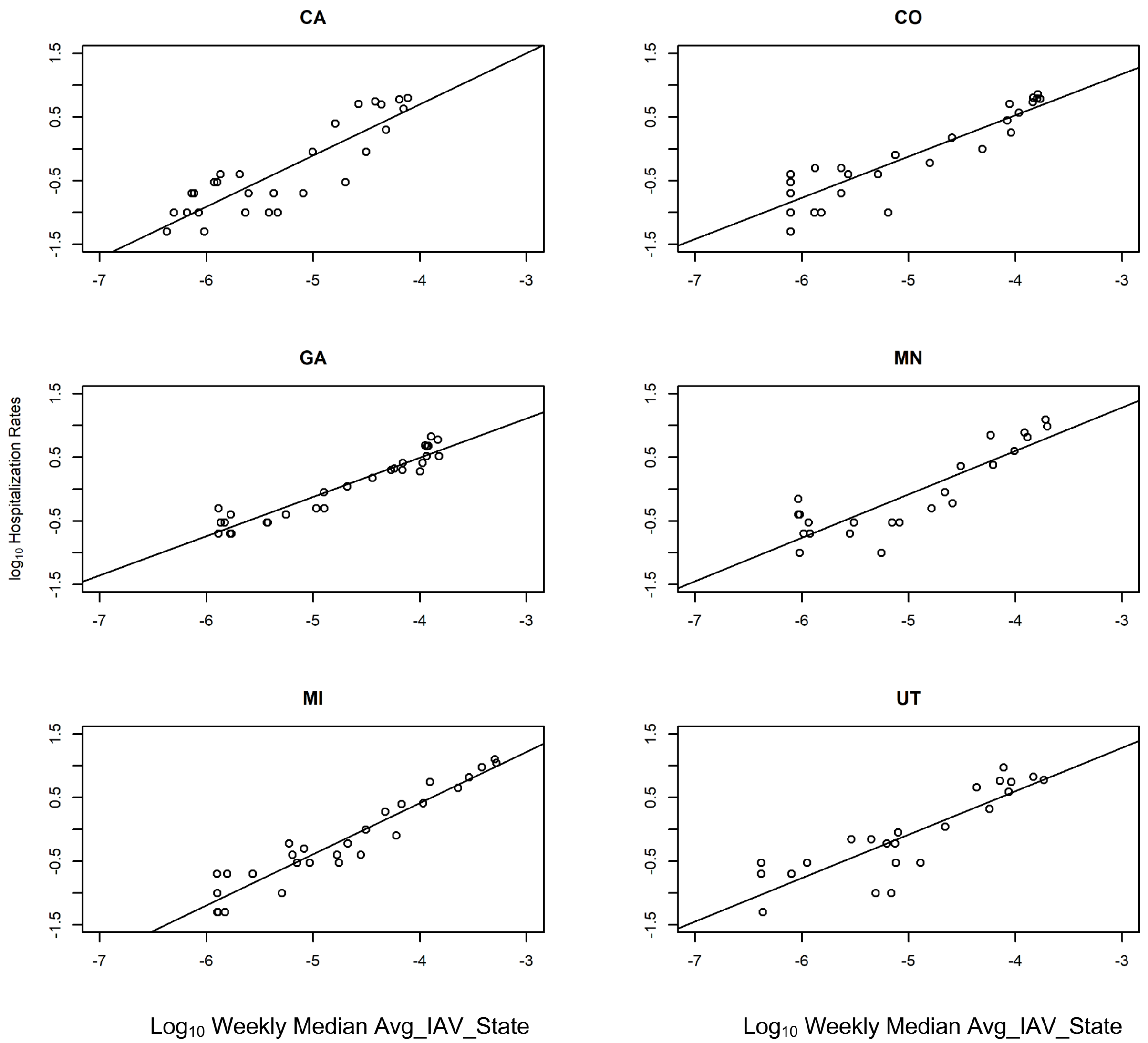

**Figure S8** Linear relationship between log10 Avg_IAV_State (expressed as median daily values each week) and Log10 hospitalization rate (reported weekly) for states with both datasets over the 2022-2023 season.

**Table S7.** Slopes, intercepts and adjusted R2 values of linear relationships in Figure S7 between log10 Avg_IAV_State (expressed as median daily values each week) and log10 hospitalization rate for selected states for 2022-2023 flu season (***p < 0.001)

| State | Slope | Intercept | Adj R2 |
|---|---|---|---|
| CA | 0.80*** | 3.91*** | 0.74 |
| CO | 0.64*** | 3.12*** | 0.81 |
| GA | 0.6*** | 2.96*** | 0.90 |
| MN | 0.68*** | 3.33*** | 0.74 |
| MI | 0.80*** | 3.63*** | 0.90 |
| UT | 0.68*** | 3.33*** | 0.72 |
| US | 0.81*** | 3.81*** | 0.89 |
